# Perturbation-Based Balance Training Reduces Falls and Fall Injuries in Older People: Insights on Mechanisms and Optimal Training Parameters from a Systematic Review and Meta-Analysis

**DOI:** 10.1101/2025.07.23.25331962

**Authors:** Shivam Sharma, Ildiko-Zsuzsa Szabo, Mathias Brix Danielsen, Stig Andersen, Jens Eg Nørgaard, Stephen R. Lord, Yoshiro Okubo, Martin Gronbech Jørgensen

**Author notes:** Correspodning Author Yoshiro Okubo, PhD.

## Abstract

Falls are common among older adults and significantly impact their quality of life. Perturbation-based balance training (PBT) is a task-specific intervention designed to improve reactive balance. This systematic review evaluated the efficacy of PBT in reducing falls, injurious falls, and fall risk factors in older adults. PubMed, CINAHL, EMBASE, and Cochrane databases were searched from inception to June 2024. Randomised controlled trials (RCTs) assessing the effects of PBT on falls or fall risk factors, including reactive balance via laboratory-induced perturbations, were included. Two reviewers independently screened studies and extracted data. Meta-analyses were conducted using a random-effects model, and certainty of evidence was assessed using the GRADE approach. Twenty-five RCTs involving 2,659 participants were included. PBT significantly reduced fall rates by 23% (rate ratio [RR]⍰0.77, 95% CI 0.60–0.99; I^2^=57%, low certainty) and injurious falls by 24% (RR⍰0.76, 95% CI 0.58–0.98; I^2^=0%) compared to controls. PBT improved reactive balance (e.g., lab-induced falls) but had limited effects on other fall risk factors (e.g., gait). In the dosage subgroup analysis, only PBT programs for at least 6 hours reduced falls by 33%. PBT reduces falls and related injuries, likely through enhanced reactive balance rather than physical improvements. While many PBT programs are low doses (∼2 hours), higher doses (6+ hours) likely offer greater fall protection. Further high-quality trials using accessible PBT methods are needed to support scalable implementation.

## INTRODUCTION

Falls in older people are a global health challenge associated with serious injuries such as hip fractures and head trauma, hospitalisations, nursing home admissions, fear of falling, poor quality of life and death (Shumway-Cook et al., 2009; World Health Organization, 2021). Age-related functional declines may be manifested as poor static balance (Lord et al., 1991), slow gait (Quach et al., 2011), decreased mobility (Beauchet et al., 2011) and poor executive function (Kearney et al., 2013) which have been the target of many conventional exercise interventions (Di Lorito et al., 2021). However, most falls in community-dwelling older people stem from unexpected perturbations such as tripping or slipping while walking (Berg et al., 1997). When such a mechanical perturbation abruptly shifts the center of mass (CoM) beyond the base of support, quick and large reactive stepping and sufficient limb support are necessary to avoid falling (Hof et al., 2005; Okubo, Schoene, et al., 2021; Patla, 2003). Reactive balance control against varied perturbations is essential for fall prevention in older adults but is not adequately addressed by conventional exercises (Kim et al., 2021).

Perturbation-based balance training (PBT), also known as reactive balance training, is a task-specific intervention designed to enhance reactive balance control through repeated exposure to postural perturbations (McCrum et al., 2022). Since its emergence in the early 2000s, various modalities of PBT have been explored, including specialised treadmills (Lurie et al., 2020; Jon D Lurie et al., 2013; Nørgaard et al., 2023), simulated slips and trips (Pai et al., 2010; Y. C. Pai et al., 2014), and manual perturbation techniques (Mansfield et al., 2018). Our previous review (2017) demonstrated that PBT could reduce by approximately 50% in older adults (Okubo et al., 2017). More recently, a review reported a 39% reduction in falls following reactive balance training (Devasahayam et al., 2022); however, this analysis included younger and disease-specific populations, potentially limiting its relevance to older adults.

As the body of evidence surrounding PBT continues to grow, with some inconsistencies emerging across trials (Aviles et al., 2019; Rogers et al., 2021; Wang et al., 2022), an updated systematic review is warranted to synthesise current findings and address unresolved clinical questions. Specifically, it remains unclear whether PBT reduces fall-related injuries and risk factors such as reactive balance and static balance, gait, mobility, and executive function. Furthermore, optimal training parameters—including dosage, perturbation type and combination with other exercises, have yet to be established, and it is unknown whether effects differ across subgroups of older adults with varying fall risk profiles.

This review aims to clarify the efficacy of PBT in reducing falls and fall-related injuries among older adults, while also evaluating its impact on key risk factors. It further explores optimal training parameters and examines whether effects differ across older adults at high versus low risk of falling, to inform clinical practice and fall-prevention guidelines.

## METHODS

### Protocol and registration

This review complies with the Preferred Reporting of Items for Systematic Review and Meta-analysis (PRISMA) (Ardern et al., 2022) in the sport and exercise medicine, musculoskeletal rehabilitation and sports science field (PERSIST) (Ardern et al., 2022). The protocol of the review was prospectively at the International Prospective Register of Systematic Reviews (PROSPERO: CRD42022343368).

### Eligibility criteria

#### Type of studies

Randomised controlled trials (RCTs) conducted in all settings were included. Only articles published in English or Japanese were included.

#### Participants

The population of interest was adults aged 60 years or older. To evaluate the effect on non-diseases specific population as seen in a municipal or open clinic training offers, we excluded studies that specifically recruited people diagnosed with neurological diseases (e.g., Parkinson’s, dementia, multiple sclerosis) or conditions (e.g., stroke).

#### Intervention

To be included in this review, studies had to include PBT in at least one intervention arm. PBT is defined *as balance training that uses repeated, externally applied mechanical perturbations to trigger rapid reactions to regain postural stability in a safe and controlled environment* (McCrum et al., 2022).

#### Comparators

All types of comparators (e.g. exercise without perturbation or no intervention) were included. However, studies were excluded if all intervention arms received PBT. If multiple comparators were reported, the most passive comparator was included as the comparator.

#### Outcome measures

The primary outcomes were fall rates, proportion of fallers, injurious falls, and laboratory-based measures of reactive balance. These outcomes were chosen because of their clinical relevance to fall prevention and frequent reporting in PBT literature. Secondary outcomes, including measures of static balance (keeping the centre of pressure (CoP) within the static base of support) (Paillard & Noé, 2015), gait speed, mobility (e.g., Timed Up-and-Go test), fear of falling, and other biomechanical markers (e.g., margin of stability), were included to explore broader effects of PBT on fall-related risk factors.

#### Duration of follow-up

No limitation on the duration of follow-up was imposed. Although if the outcome is assessed at multiple time points, the one closest to 12 months was used.

#### Information sources and search strategy

Systematic searches were performed in four electronic databases (PubMed, CINAHL, EMBASE, and Cochrane) from inception to June 2023. An updated search was conducted in July 2024. The search string was built with a research librarian using key terms for population, intervention, and study design (clinical trials, RCTs). No language restriction was imposed during the data-base search. Reference lists of included studies and relevant systematic reviews were also manually searched, and experts in the field were contacted to identify any missing studies. Search strategy is available in supplementary material.

#### Study selection

Two reviewers (ISZ, JEN/SS) independently screened abstracts and titles based on the eligibility criteria. The reviewers (ISZ, JEN/SS) also assessed the full text of the retrieved studies to determine inclusion.

Conflicting opinions were solved by discussions between reviewers and, if necessary, by consulting a third reviewer (MGJ/YO). The study selection process was conducted in RAYYAN (Ouzzani et al., 2016) and Covidence (for updated searches).

#### Data collection process

Two reviewers (ISZ, SS) independently extracted data in Covidence using a template covering study design, participants, interventions, outcomes, and results. Disagreements were resolved through discussion or additional reviewers (MGJ, YO). List of extracted data items is available in the supplementary material.

#### Risk of bias assessment

Two authors (SS, MBD) individually assessed risk of bias in the included studies using the Cochrane risk of bias tool 2 (ROB 2) (Sterne et al., 2019). All outcomes were separately assessed using ROB 2.

Disagreements during the consensus process were resolved through mutual discussion between reviewers (YO, MGJ), with final decisions made following appropriate justification. When available, trial protocols and registrations were retrieved and reviewed to supplement information provided in the included studies.

#### Strategy for data analysis

The results from studies with sufficiently similar methodologies were combined in a series of meta-analyses using DerSimonian and Laird inverse variant random effect model in Cochrane Review Manager software (RevMan 5.0). Fall rates were reported as incidence rate ratios (IRRs) with their corresponding 95% confidence intervals (CIs). Binary outcomes, including the proportion of fallers, injurious fallers, and laboratory fallers, were presented as risk ratios (RRs) with 95% CIs. For continuous variables, such as margin of stability, hip height, step height, fear of falling, mobility, static balance and gait speed, results were reported as mean differences or standardised mean differences with their respective 95% CIs. Statistical heterogeneity was determined by visual inspection of forest plots and I^2^ statistics (≥75%; considerable heterogeneity).

Planned subgroup analyses were conducted to examine the differences in fall rates by (i) participant characteristics (higher risk of falling vs lower risk of falling) (e.g. based on fall history, balance impairment), (ii) PBT type (walkway vs treadmill vs manual/standing perturbation), (iii) training dosage (≤ 6 hours vs >6 hours), and (iv) intervention type (PBT alone vs combined PBT, such as PBT plus resistance training) and (v) exercise without perturbation vs no intervention compared with PBT as common comparator. We chose these subgroups because prior evidence suggests effectiveness varies by dosage, perturbation type, and risk profile.

Sensitivity analyses were performed comparing the results of the low-risk bias studies with the high-risk bias studies. Leave-one-out analyses were performed to assess the robustness of our findings on fall rates and to evaluate whether the exclusion of individual studies significantly altered the overall effect size or heterogeneity.

#### Publication bias

Publication bias was assessed by visually inspecting funnel plots to identify potential asymmetry, which could indicate selective reporting of results.

#### Grading of Evidence

The certainty of evidence was evaluated using the GRADEpro Guideline Development Tool (Table 1) (McMaster University, 2023). The overall quality of evidence was determined as high (further research is very unlikely to change our confidence in the estimate of effect), moderate (further research is likely to have an important impact on our confidence in the estimate of effect and may change the estimate), low (further research is very likely to have an important impact on our confidence in the estimate of effect and is likely to change the estimate), or very low (any estimate of effect is very uncertain).

## RESULTS

### Study Selection

The initial search identified 1932 articles after removal of duplicates. Of these, 53 articles were retrieved for full text assessment, and 23 were identified as meeting all criteria and were thus considered eligible for inclusion in our review. An additional updated search was conducted in July 2024 to include any new studies published while the review was underway. The updated search revealed 144 new studies after the removal of duplicates, and after full-text screening, two studies met the inclusion criteria. Consequently, 25 studies were deemed eligible for inclusion after the full-text screening.

### Study Characteristics

Of the 25 RCTs included in this review, six used perturbation walkaways (e.g. sliding tiles, pop-up obstacles) (Allin et al., 2020) (Bhatt et al., 2021; Liu et al., 2021; Y Okubo et al., 2019; Y.-C. Pai et al., 2014; Rogers et al., 2021), 16 used treadmills (e.g. belt accelerations/decelerations)(Aviles et al., 2019; Bieryla et al., 2007; Brull et al., 2023; Gerards et al., 2023; Gimmon et al., 2018; Kurz et al., 2016; Lurie et al., 2020; J. D. Lurie et al., 2013; Nørgaard et al., 2023; Obuchi et al., 2004; Rieger et al., 2020; Shimada et al., 2004; Wang et al., 2019; Wang et al., 2022) and four used manual/standing perturbations (e.g. surface translation) (Maki et al., 2008; Mansfield et al., 2010; Parijat & Lockhart, 2012; Wang et al., 2022) as their PBT modality. Out of 25 studies only one study had no intervention for the control group (Brull et al., 2023). Rest all other studies had non-PBT exercises. Training doses were >6 hours (average 9.8 hours) in seven studies (Aviles et al., 2019; Brull et al., 2023; Gimmon et al., 2018; Kurz et al., 2016; Maki et al., 2008; Rogers et al., 2021; Shimada et al., 2004), and ≤6 hours (average 1.6 hours) in the remaining included studies. Eight studies (Brull et al., 2023; Gerards et al., 2023; Lurie et al., 2020; Jon D Lurie et al., 2013; Maki et al., 2008; Mansfield et al., 2010; Rieger et al., 2024; Shimada et al., 2004) included older people at increased risk of falls while the other studies included general samples of older people. A detailed methodology of these RCTs is reported in Supplementary Table 1. The intensity of perturbation was reported inconsistently and often inadequately across studies, with treadmill belt accelerations ranging from 0.1 to 20 m/s, and moveable platform slip distances varying from 10 cm to 90 cm. Only one study (Rieger et al., 2024) reported the use of harness-supported falls during training.

### Results of syntheses

#### Effect of PBT intervention on fall outcomes

Eleven trials (n=1231) included data on total falls (rate ratio) and fallers (risk ratio). Meta-analyses of these trials showed significant reductions in the rate of falls (Figure 1; rate ratio: 0.77, 95% CI: 0.60 to 0.99, p=0.040, *I^2^* =57%) and proportion of fallers (Figure 2; risk ratio=0.83, 95% CI 0.74 to 0.93, p=0.001, *I^2^* =0). A significant reduction in injurious falls was also observed across four studies, with a pooled risk ratio of 0.76 (Figure 3; risk ratio=0.76, 95% CI: 0.58 to 0.98, p=0.030, *I^2^* =0%).

#### Effect of PBT intervention on fall-related risk factors

Laboratory-assessed falls related to reactive balance demonstrated a significant reduction following PBT intervention across nine trials (Figure 4; risk ratio =0.42, 95% CI: 0.31 to 0.58, p<0.001, *I^2^* =38%). There were also improvements in margin of stability (5 studies, n =381, MD (m)=0.47, 95% CI: −0.01 to 0.94, *p* =0.050, *I^2^* =68%) and hip height (4 studies, n =206, SMD (m)=0.43, 95% CI: 0.05 to 0.940, *p* =0.05, *I^2^* =68%).

Secondary fall-related risk factors including fear of falling (6 studies, *n* =579), gait speed (5 studies, *n* =126), static balance (4 studies, *n* =527), step length (8 studies, *n* =288), and performance on the Timed Up and Go (TUG) test (5 studies, *n* =418) were analysed. However, none of these outcomes demonstrated statistically significant improvements following PBT intervention compared to control groups. Individual forest plots for each factor are presented in the supplementary appendices. Eight studies reported adverse events during training (Brull et al., 2023; Gerards et al., 2017; Kurz et al., 2016; Lurie et al., 2020; Mansfield et al., 2010; Nørgaard et al., 2023; Y Okubo et al., 2019; Y. Okubo et al., 2019; Rogers et al., 2021) but other 17 studies did not.

#### Subgroup analyses

Since not all included studies reported all secondary outcomes, only studies that reported the outcome of interest were included in the meta-analysis. Forest plots for each subgroup analysis are provided in the online supplementary. Regarding combination, PBT combined with exercise significantly reduced falls (rate ratio = 0.26; 95% CI: 0.13 to 0.54, *p* < 0.001) but not PBT alone (rate ratio = 0.81; 95% CI: 0.62 to 1.06; *p* = 0.120). The falls reduction was significantly greater for PBT combined with exercise (*p* for subgroup = 0.004). When stratified by training dosage (<6 hours vs 6+ hours), subgroup difference was not significant (*p* for subgroup = 0.46), but only PBT with higher training dose (>6 hours) significantly reduced falls (rate ratio = 0.67; 95% CI: 0.46 to 0.97; *p* = 0.03). Similarly, no significant subgroup differences were observed in participant characteristics (healthy vs high-risk, *p* = 0.79) and PBT type (walkway vs treadmill vs manual/standing; *p* = 0.27) but only walkway PBT significantly reduced falls (rate ratio = 0.50; 95% CI = 0.28, 0.9; *p* = 0.02). Regarding the control type, no significant subgroup difference was observed between trials comparing PBT against exercise without perturbations, and those comparing PBT against no exercise (*p* for subgroup = 0.10).

#### Sensitivity analyses

Sensitivity analysis comparing studies with low versus high risk of bias revealed no significant difference in effect size for the total number of falls (*p* = 1.00; I = 0%). Influence analysis using a leave-one-out approach for daily falls indicated that the overall effect size became non-significant when certain studies (Jon D Lurie et al., 2013; Nørgaard et al., 2023; Y. C. Pai et al., 2014; Rieger et al., 2024; Rogers et al., 2021; Shimada et al., 2004) were excluded, with heterogeneity (I^2^) ranging from 44% to 59%. Notably, heterogeneity was substantially reduced from 55% to 16% when the study by Lurie et al. (2020) (Lurie et al., 2020) was omitted (*p* = 0.050).

#### Publication bias

Due to the limited number of included studies, the funnel plots for total number of falls, number of fallers, and injurious falls lacked a discernible funnel shape, preventing meaningful interpretation (supplementary file). However, the funnel plot for laboratory falls indicated potential publication bias. Upon removal of suspected outlier studies (Liu et al., 2015; Liu et al., 2021; Parijat & Lockhart, 2012), the effect size was no longer statistically significant, suggesting that these studies may have disproportionately influenced the original findings.

#### Risk of bias (ROB) and certainty of evidence

The GRADE assessment indicated that the certainty of evidence was low for outcomes related to risk of falls, injurious falls, and moderate for the risk of fallers and laboratory-assessed falls. These ratings were primarily influenced by methodological limitations across the included studies, such as lack of concealed allocation, absence of blinding procedures, potential selective outcome reporting, and incomplete outcome data due to attrition.

## DISCUSSION

This review revealed PBT reduced fall rates in older adults by 23% compared to control groups receiving exercise without perturbation or no exercise/intervention. However, the certainty of this evidence was rated as low. Similarly, PBT was associated with a 24% reduction in injurious falls, also supported by low-certainty evidence. In contrast, the proportion of older adults experiencing at least one fall decreased by 17%, with moderate certainty in the estimate. PBT enhanced reactive balance responses to sudden perturbations. PBT did not significantly affect static balance, gait speed, or fear of falling, suggesting that fall reduction might stem from improved reactive balance rather than broader physiological or psychological changes.

### Mechanisms

PBT differs fundamentally from traditional balance training. (Sherrington et al., 2019; Sherrington et al., 2017; Wang et al., 2022) While conventional exercises often focus on maintaining the center of mass within a reduced base of support (e.g., single-leg stance), PBT targets reactive balance, i.e.: the ability to recover from sudden, unexpected perturbations. This is a critical mechanism of fall prevention that conventional programs do not adequately address. (Grabiner et al., 2014; McCrum et al., 2022) PBT achieves this by exposing individuals to repeated, controlled perturbations, allowing them to develop predictive and reactive motor strategies to maintain balance.

### Improved Reactive Balance and Transfer to Daily Life

The review found that PBT was associated with a 58% reduction in laboratory-induced falls, suggesting a improvement in reactive balance. Further, reduction in real-world falls was more dampened at approximately 25%. This difference may reflect the complexity and unpredictability of real-life environments, which current PBT protocols may not fully replicate (Phu et al., 2024; Song et al., 2021). Moreover, many PBT programs are delivered over short timeframes, and their protective effects may diminish over time. For instance, one trial reported a 57% reduction in injurious falls at three months follow-up, but this benefit was lost at 12 months (Lurie et al., 2020). These findings highlight the potential value of booster sessions to maintain long-term benefits (Bhatt et al., 2012).

Biomechanical outcomes also support the efficacy of PBT. Improvements were observed in margin of stability and hip height, suggesting enhanced postural control. However, changes in other measures, such as recovery step length, were not significant, likely due to variability in perturbation types (e.g., slips vs. trips), which elicit different motor adaptations (Phu et al., 2024). This underscores the need for more standardised outcome measures in future studies.

### Dose and intensity of PBT

Interestingly, most PBT programs included in this review involved relatively low training doses, averaging just 1.6 hours across 3–6 sessions. This suggests, PBT may reduce falls through motor learning, enabling quick gait and posture adaptation. Studies show that older adults often adjust within 3–24 trials in a single session using predictive strategies like forward center-of-mass shifts (Bohm et al., 2015; Pai et al., 2010; Y. C. Pai et al., 2014). However, in the dosage subgroup analysis, only PBT programs with at least 6 hours reduced falls by 33%. While optimal training dose may vary for each individual, several hours of PBT may be necessary to develop familiarity with perturbations, learn effective reactive balance strategies, and automate these responses (Okubo et al., 2018; Y Okubo et al., 2019), especially in frailer individuals who benefit from gradual progression starting from lower-intensity perturbations. However, six hours are still substantially lower than the recommended dose for conventional exercises (3 hours/week on an ongoing basis) (Sherrington et al., 2017).

Due to the inherent heterogeneity of PBT methods, the intensity of interventions has been reported inconsistently and lacks reproducibility across studies. We recommend that future research consistently report the number and rate of harness-supported falls as a universal and objective indicator of PBT intensity (Rieger et al., 2024). Incorporating such measures may facilitate tailored interventions and enhance clinical outcomes.

### Different PBT Methods

Subgroup analyses did not reveal statistically significant differences between PBT delivery methods—such as treadmills, walkways, or manual/standing perturbations. However, walkway interventions showed a larger effect size, suggesting a potential trend toward greater efficacy. Some studies report generalisation of skills from treadmill to overground settings, while others highlight distinct adaptations and limited transferability (Lee et al., 2018; Phu et al., 2024; Song et al., 2021). These Mixed findings question whether all PBT modalities are equally effective, underscoring the need to identify methods that best mimic real-world perturbations.

### Complementing Conventional Exercise

Since PBT did not significantly improve other risk factors, balance, gait speed, or fear of falling, in our opinion it should be considered a complementary intervention rather than a substitute for conventional exercise. Notably, subgroup analysis based on one study revealed that combining PBT with lower-limb strength training reduced falls by 74%, a significantly greater effect than PBT alone. This supports the idea that muscle strength, particularly in the hips, is essential for executing rapid reactive steps (Okubo, Duran, et al., 2021; Rogers et al., 2021). As it is a single-study analysis, the conclusion remains uncertain and future research should focus on this approach. For now, we suggest integrating PBT with traditional strength and balance exercises, as this combination may offer synergistic benefits and address multiple fall risk factors. Additionally, there was no significant subgroup difference when comparing exercise without perturbation and no exercise against PBT. However, it is noteworthy that PBT reduced falls by 45% compared to no exercise (*p* = 0.02), but only by 13% compared to exercise without PBT (*p* = 0.24).

### Clinical Implications

PBT effectively improves reactive balance and should be integrated into fall prevention programs for both healthy and high-risk older adults. Brief PBT sessions can provide benefit, but programs should aim for higher doses (≥6 hours) when feasible to achieve sustainable neuromuscular adaptations. Combining PBT with exercise might offer synergistic benefits. Manual perturbations are practical and low-cost for widespread implementation, while walking-based perturbations may enhance generalisability to real-life situations when resources allow. However, given the low certainty of the current evidence, more rigorous high-quality trials are needed.

### Limitations

Despite its strengths, this review has several limitations. First, the possibility of publication bias and missed studies in including those not published in English or Japanese cannot be ruled out. Second, there was substantial heterogeneity in PBT protocols, including differences in training modality, dosage, and delivery, which may have influenced the pooled results. Third, although sensitivity and subgroup analyses were conducted, these one-dimensional approaches cannot fully account for the complex confounding present across studies. Fourth, moderate heterogeneity (I = 55%) was observed in the meta-analysis, which was reduced to 16% upon excluding Lurie et al. (2020) (Lurie et al., 2020), indicating it may have disproportionately contributed to between-study variability, likely due to its pragmatic design and lack of a standardized PBT protocol (e.g., session range: 1–25). Lastly, the limited number of studies included and their methodological weaknesses, resulted in low to moderate certainty of evidence. These limitations highlight the need for high-quality, standardised trials to strengthen the evidence base and clarify optimal PBT practices.

## Conclusion

This review provides evidence that PBT reduces all fall rates in older adults compared to exercises without perturbations or no exercise. PBT clearly improves reactive balance control, as indicated by a 58% reduction in laboratory-induced falls and improved dynamic stability following perturbations. Low-dose PBT (1.5 hours) likely promotes rapid motor learning and anticipatory strategies, whereas high-dose PBT (>6 hrs) may drive more sustainable, generalizable neuromuscular adaptations for fall prevention.

Combining PBT with exercise may offer synergistic benefits, as PBT alone may not improve other risk factors. Evidence for delivery methods is mixed: fall reduction was demonstrated clearly in walkway-based PBT, while differences from treadmill-based and manual PBT remain inconclusive. Thus, manual/standing perturbation techniques remain a feasible, low-cost option for clinical implementation.

Overall, PBT is a clinically relevant and effective intervention that can be integrated into routine exercise programs to strengthen fall prevention strategies in older adults. Given the limited evidence, high-quality trials using accessible and scalable PBT delivery methods are needed to confirm its effectiveness for widespread community implementation.

## Contributors

All authors contributed to the development of the study protocol and design. IZS and MGJ performed the literature searches. IZB and JEN screened titles, abstracts, and full-text articles. SS, MBD, YO, and MGJ were responsible for data extraction and risk of bias assessment. Certainty of evidence was assessed by MGJ and YO. SS and YO conducted the data analysis and drafted the initial manuscript. YO served as the study guarantor. SRL and SA reviewed and approved the final manuscript after comments of all authors.

## Patient involvement and consent

Not applicable.

## Ethics approval

Not applicable.

## Equity, diversity and inclusion (EDI) statement

Not applicable.

## Data sharing statement

All data relevant to this study are included within the article and supplementary material. PROSPERO Registration: CRD42022343368

## Data Availability

All data produced in the present study are available upon reasonable request to the authors

## FIGURE LEGENDS

**Figure 1** Result of meta-analyses for rate ratio of falls (Rogers 2021a – induced step training plus hip strengthening vs hip strengthening; Rogers 2021b – induced step training vs flexibility and relaxation). Risk of bias (A) Randomisation process, (B) Deviations from the intended interventions, (C) Missing outcome data, (D), Measurement of the outcome, (E) Selection of the reported result, (F) Overall

**Figure 2** Result of meta-analyses for risk ratio (fallers) (Rogers 2021a – induced step training plus hip strengthening vs hip strengthening; Rogers 2021b – induced step training vs flexibility and relaxation). Risk of bias (A) Randomisation process, (B) Deviations from the intended interventions, (C) Missing outcome data, (D), Measurement of the outcome, (E) Selection of the reported result, (F) Overall

**Figure 3** Result of meta-analyses for risk ratio (Injurious falls) Risk of bias (A) Randomisation process, (B) Deviations from the intended interventions, (C) Missing outcome data, (D), Measurement of the outcome, (E) Selection of the reported result, (F) Overall

**Figure 4** Result of meta-analyses for risk ratio (laboratory fallers) (Liu 2020a - overground slip-perturbation training vs Ot: overground slip-perturbation training; Liu 2020b – treadmill perturbation training vs treadmill control; Allins 2020a – slip training vs control). Risk of bias (A) Randomisation process, (B) Deviations from the intended interventions, (C) Missing outcome data, (D), Measurement of the outcome, (E) Selection of the reported result, (F) Overall

---

**Table 2.**
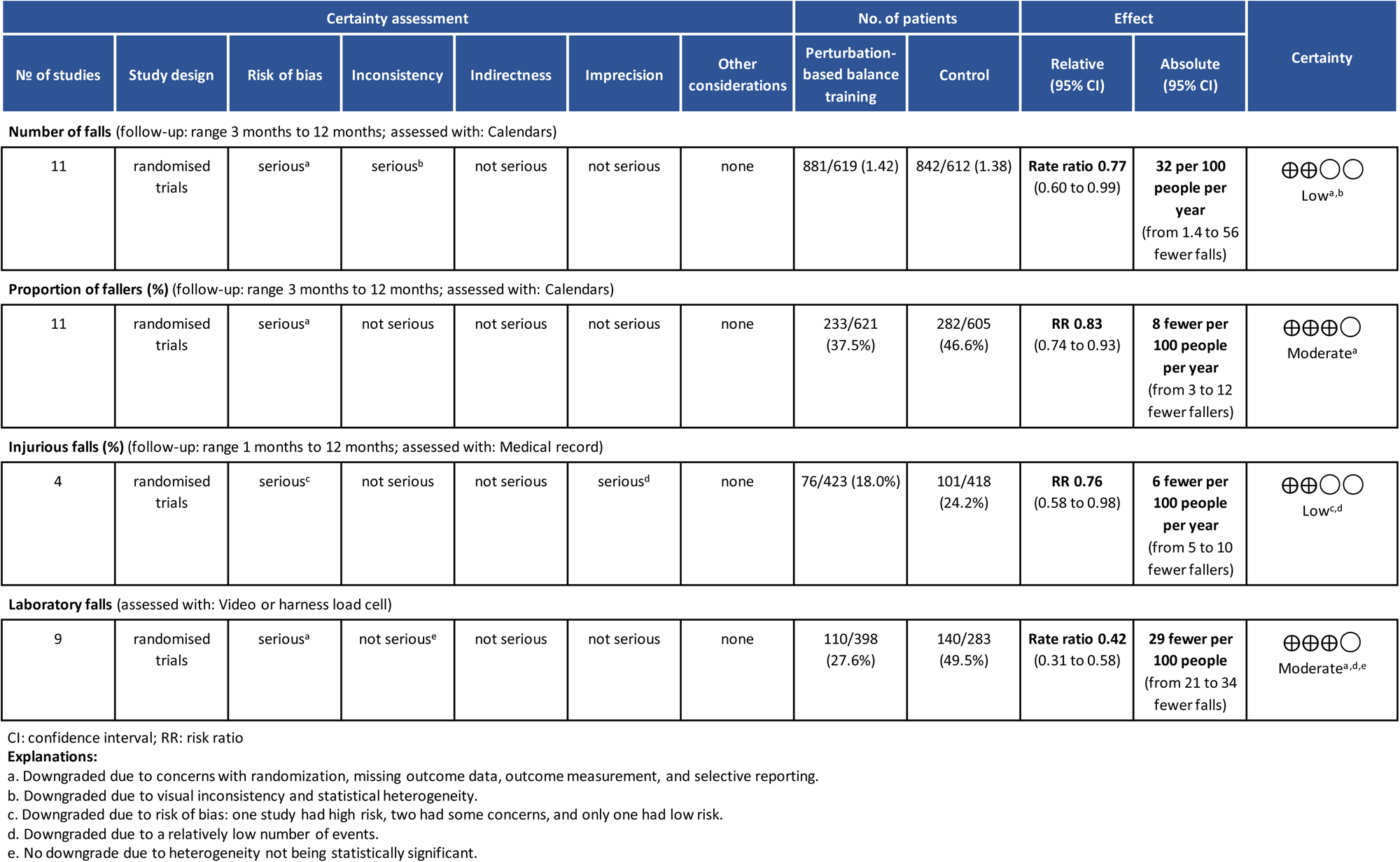
Results of GRADE evaluation.

## Supplementary material 1: SEARCH HISTORY

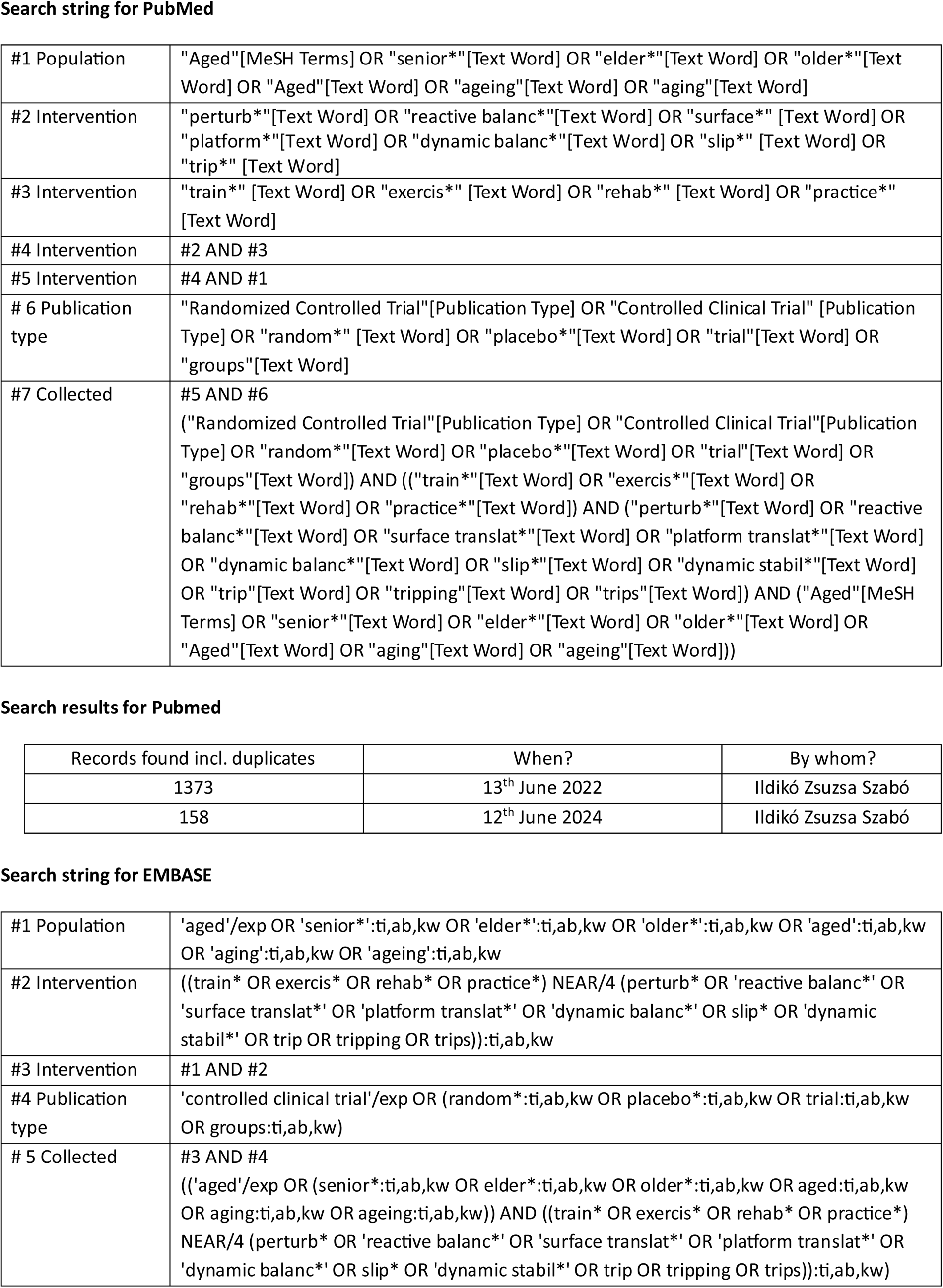
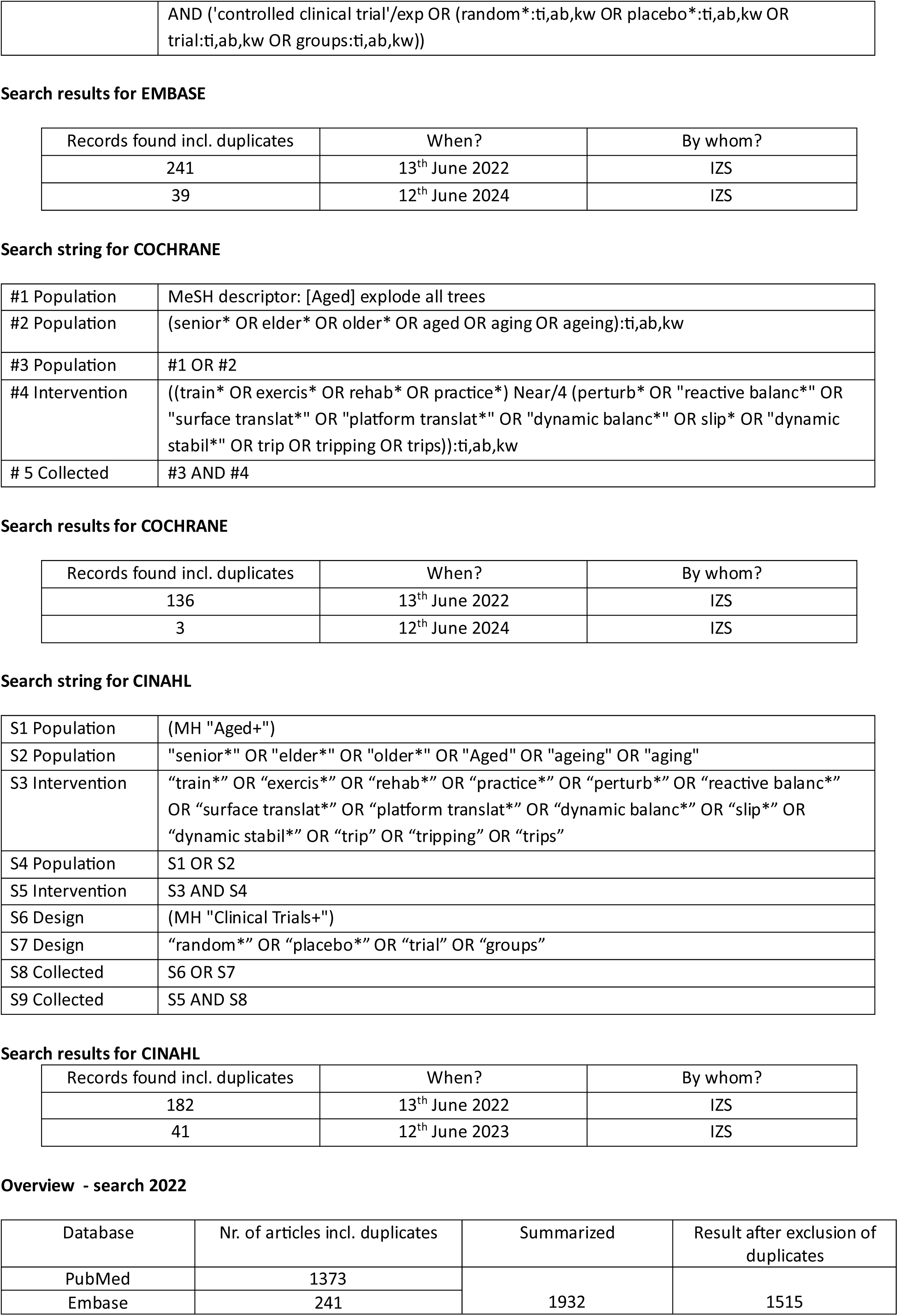
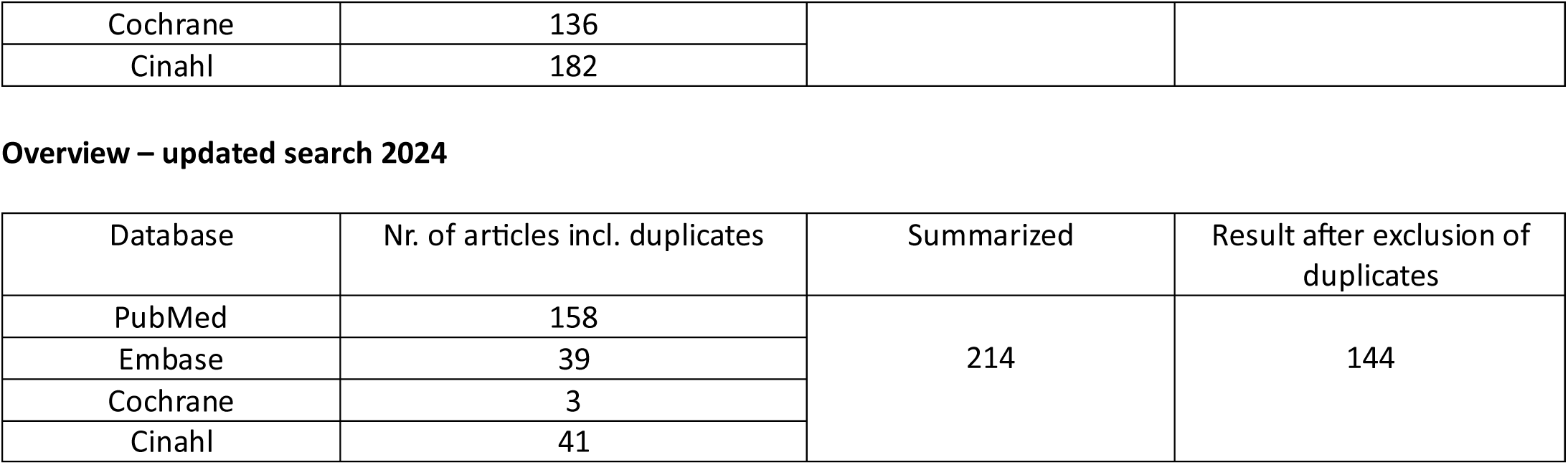
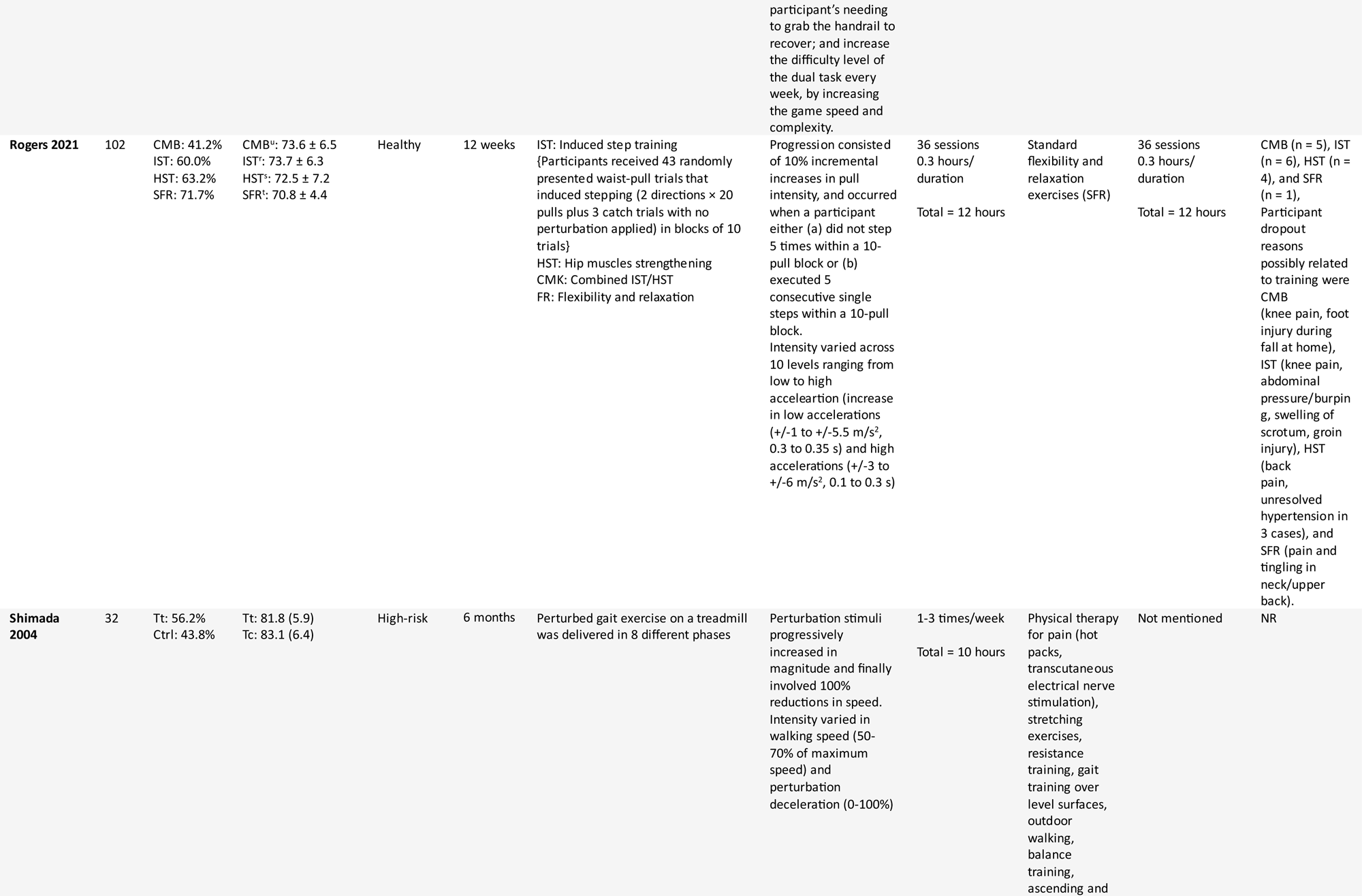

## Supplementary material 2: PRIORITIZED LIST FOR TITLE AND ABSTRACT

| Study design |  |
| --- | --- |
| Inclusion | Exclusion |
| 1. Randomised, controlled trials<br>2. Cluster Randomised, controlled trials | 1. Systematic review and meta-analysis<br>2. Cohort Study<br>3. Case-control study<br>4. Cross-sectional study<br>5. Case reports or series |
| Population |  |
| Inclusion | Exclusion |
| 1. $\geq 60$ years<br>2. Community-dwelling<br>3. No information about location of recruitment and administration of intervention<br>4. Participants with increased risk of falling<br>a. e.g. previous fallers, mild cognitive impairment<br>5. No information about conditions or diseases | 1. If specific conditions or diseases is stated as an inclusion criteria<br>a. e.g. Parkinson's, stroke, multiple sclerosis, osteoporosis)<br>2. If participants are in-patients at hospitals, departments, or wards<br>3. If participants are enrolled at nursing or residential care facilities |
| Intervention |  |
| Inclusion | Exclusion |
| 1. Gait adaptability training<br>a. Definition; "training consisting of voluntary gait and/or stepping that is dictated by the cognitive processing of unpredictable sensory stimuli"<br>i. Music-based multitask training (e.g. Dalcroze Eurhythmics)<br>ii. Obstacle course training<br>iii. Target stepping training<br>iv. Virtual-reality step and/or gait training<br>v. Step or gait-based exergaming<br>2. Multimodal exercises will be included<br>a. Studies were gait adaptability training is combined with another training modality | b. Strength/resistance training<br>c. Balance training<br>d. Endurance training<br>e. General physical activity<br>f. Dual-task training with unrelated cognitive-task<br>b. Walking while during verbal fluency, arithmetic or likewise.<br>g. Flexibility training<br>h. 3D interventions<br>a. e.g. dance, tai-chi, Qi gong |
| Comparator |  |
| Inclusion | Exclusion |
| All comparators will be included | Not applicable |
| Outcomes |  |
| Inclusion | Exclusion |
| 1. Fall data collected prospectively for at least 6 months | 1. No fall data is collected.<br>2. Fall data is collected for less than 6 months. |

## Supplementary material 3: DATA ITEMS EXTRACTED

| General information |  |
| --- | --- |
| Author(s) | Report the lead author and full name list |
| Publication year | Report publication year |
| Title of study | Report title of study |
| Protocol registry | Report registry name |
| Protocol registry number | Report registry number |
| Publication type | Journal article/Abstract/Other: elaborate |
| Years of study | Report period of time the study was conducted |
| Country of study | Report where the study was conducted |
| Funding source of study | Report source of study |
| Potential conflict of interest from funding | Yes/None declared/No/Unclear |
| Type of study | Individually randomized, controlled trial/Cluster randomized, controlled trial/Pilot controlled trial/Other: elaborate |
| Primary aim of study | The effect on falls/Effect on fall-related risk factors/Not clear/Other: elaborate |
| Participant characteristics |  |
| Inclusion and exclusion criteria | Report criteria for participation and exclusion stated in the study? |
| Participant setting | Report the residential status of the participants |
| Participant health status | Report if increased risk of falling or not |
| Age | Mean (SD or SE) or age range of included participants |
| Gender | Report %-female participants |
| Fall history | Report proportion of the population that have had prior falls |
| Participants randomized | Report total, intervention group and control group |
| Number of dropouts | Report total, intervention group and control group |
| Intervention characteristics |  |
| Intervention description | Report the perturbation-based balance training protocol |
| Category of perturbation used | Overground walkways with movable platform and tripboards/specialized treadmills that produce well-timed accelerations/treadmills with ankle-mounted brake-and-release system/therapist-applied perturbations/movable platforms that make sudden movements/other: elaborate |
| Multimodal intervention? | Yes/no |
| Intervention setting | Report where the intervention was conducted |
| Supervised training | Yes/no/unclear |
| Training duration | Report length of training session |
| Training sessions per week | Report number of training sessions per week |
| Intervention dose | Report duration of intervention in weeks |
| Intervention duration | Report duration of intervention in weeks |
| Control description | Report the comparator used |
| Control training duration | Report length of comparator training session |
| Control training sessions per week | Report number of comparator training sessions per week |
| Control intervention dose | Report duration of intervention in weeks |
| Outcome measures |  |
| Main outcome: Daily life fall and fall rate (fall per person-year) | Report primary outcome measures assessed in the study:<br>- definition of fall |
|  | - method of fall assessment (fall calendar/retrospective recall of falls/unclear/other: elaborate)<br>- report months of data collected<br>- is fall rate reported?<br>- is proportion of fallers reported?<br>- is proportion of fall-related injuries reported?<br>- is proportion of fall-related fractures reported?<br>- is proportion of healthcare-contact reported? |
| **Additional outcomes: Laboratory fall** | Report laboratory fall outcome measures assessed in the study:<br>- does the study report laboratory fall metrics?<br>- how is laboratory fall defined in the study?<br>- method of laboratory fall assessment (Percentage of body weight supported in harness/Video review/Other: elaborate)<br>- time of assessment (pre-intervention/post-intervention/follow-up/not clear)<br>- report months of data collected<br>- is rate of laboratory falls reported in the study?<br>- is proportion of laboratory falls reported in the study? |
| **Additional outcomes: Reactive balance** | Report reactive balance outcome measures assessed in the study:<br>- does the study report laboratory fall metrics?<br>- method of reactive balance assessment (Margin of stability/Hip height/Step length/Other: elaborate)<br>- time of assessment (pre-intervention/post-intervention/follow-up/not clear)<br>- report months of data collected<br>- is the reactive balance following laboratory induced slips assessed in the study?<br>- is the reactive balance following laboratory induced trips assessed in the study? |
| **Additional outcomes: Fall-related risk factors**<br>(including gait pattern, balance, mobility, executive function, health-related quality of life and fear of falling) | Report fall-related risk factors outcome measures assessed in the study. For all elements included:<br>- does the study report these measures?<br>- describe how was the outcome measured<br>- time of assessment (pre-intervention/post-intervention/follow-up/not clear) |
| **Results** |  |
| **Main outcome: Daily life fall and fall rate** | Report the results regarding daily life fall and fall rate (falls per person-year); incidence rate ratio; p-values |
| **Additional outcomes: Laboratory fall** | Report the results regarding laboratory fall |
| **Additional outcomes: Reactive balance** | Report the results regarding reactive balance |
| **Additional outcomes: Fall-related risk factors** | Report the results of gait pattern, balance, mobility, executive function, health-related quality of life and fear of falling; pre- and post-scores (SD/SE) and difference (SD/SE). |

## Supplementary material 4: Leave one out meta-analysis

Supplementary material 4: Leave one out meta-analysis Daily falls (rate ratio)
| Omitted study | Effect size with 95% CI | p-value | I<sup>2</sup> (heterogeneity) % |
| --- | --- | --- | --- |
| Original | 0.77 [0.59, 0.99] | **0.04** | **57** |
| Aviles 2019 | 0.77 [0.59, 1.00] | 0.05 | 47 |
| Lurie 2020 | 0.71 [0.56, 0.90] | 0.005 | 19 |
| Pai 2014 | 0.82 [0.64, 1.05] | 0.12 | 50 |
| Shimada 2004 | 0.79 [0.61, 1.03] | 0.07 | 57 |
| Lurie, 2013 | 0.83 [0.66, 1.06] | 0.11 | 47 |
| Maki 2008 | 0.76 [0.58, 0.99] | 0.04 | 61 |
| Mansfield, 2010 | 0.75 [0.57, 0.98] | 0.03 | 60 |
| Rogers, 2021a | 0.76 [0.58, 1.00] | 0.05 | 60 |
| Rogers, 2021b | 0.81 [0.63, 1.04] | 0.11 | 49 |
| Wang 2022 | 0.72 [0.54, 0.96] | 0.02 | 60 |
| Norgaard, 2023 | 0.75 [0.56, 1.02] | 0.07 | 57 |
| Rieger 2023 | 0.77 [0.58, 1.01] | 0.05 | 60 |

Fallers (risk ratio)
| Omitted study | Effect size with 95% CI | p-value | I<sup>2</sup> (heterogeneity) % |
| --- | --- | --- | --- |
| Original | 0.86 [0.78, 0.96] | **0.001** | **0** |
| Aviles 2019 | 0.86 [0.78, 0.96] | 0.001 | 0 |
| Lurie 2020 | 0.87 [0.73, 1.03] | 0.11 | 0 |
| Pai 2014 | 0.88 [0.79, 0.98] | 0.05 | 0 |
| Shimada 2004 | 0.87 [0.78, 0.97] | 0.02 | 0 |
| Lurie, 2013 | 0.87 [0.78, 0.97] | 0.02 | 0 |
| Maki 2008 | 0.87 [0.78, 0.96] | 0.002 | 0 |
| Mansfield, 2010 | 0.86 [0.77, 0.96] | 0.001 | 0 |
| Rogers, 2021a | 0.86 [0.77, 0.96] | 0.001 | 0 |
| Rogers, 2021b | 0.85 [0.76, 0.95] | 0.003 | 0 |
| Wang 2022 | 0.86 [0.77, 0.96] | 0.0008 | 0 |
| Norgaard, 2023 | 0.86 [0.77, 0.97] | 0.002 | 0 |
| Rieger 2023 | 0.87 [0.78, 0.96] | 0.001 | 0 |

Laboratory falls (risk ratio)
| Omitted study | Effect size with 95% CI | p-value | I<sup>2</sup> (heterogeneity) % |
| --- | --- | --- | --- |
| Original | 0.42 [0.31, 0.58] | **<0.00001** | **38** |
| Allins, 2020a | 0.42 [0.30, 0.60] | **<0.00001** | 45 |
| Bhatt, 2021 | 0.37 [0.26, 0.54] | **<0.00001** | 16 |
| Liu 2015 | 0.43 [0.31, 0.59] | **<0.00001** | 42 |
| Liu 2020 a | 0.43 [0.32, 0.59] | **<0.00001** | 32 |
| Liu 2020 b | 0.41 [0.28, 0.60] | **<0.00001** | 45 |
| Wang, 2019 | 0.54 [0.46, 0.63] | **<0.00001** | 12 |
| Parijat 2012 | 0.40 [0.28, 0.58] | **<0.00001** | 32 |
| Norgaard, 2023 | 0.41 [0.28, 0.60] | **<0.00001** | 39 |
| Okubo, 2019 | 0.38 [0.26, 0.56] | **<0.00001** | 43 |

Injurious falls
| Omitted study | Effect size with 95% CI | p-value | I<sup>2</sup> (heterogeneity) % |
| --- | --- | --- | --- |
| Original | **0.76 [0.59, 0.98]** | **0.03** | **0** |
| Lurie, 2020 | 0.64 [0.41, 1.00] | 0.05 | 0 |
| **Pai, 2014** | 0.77 [0.59, 1.01] | 0.06 | 0 |
| **Norgaard, 2023** | 0.77 [0.57, 1.04] | 0.08 | 0 |
| **Lurie, 2013** | 0.77 [0.59, 1.00] | 0.05 | 0 |

---

**Supplementary Table 1.**
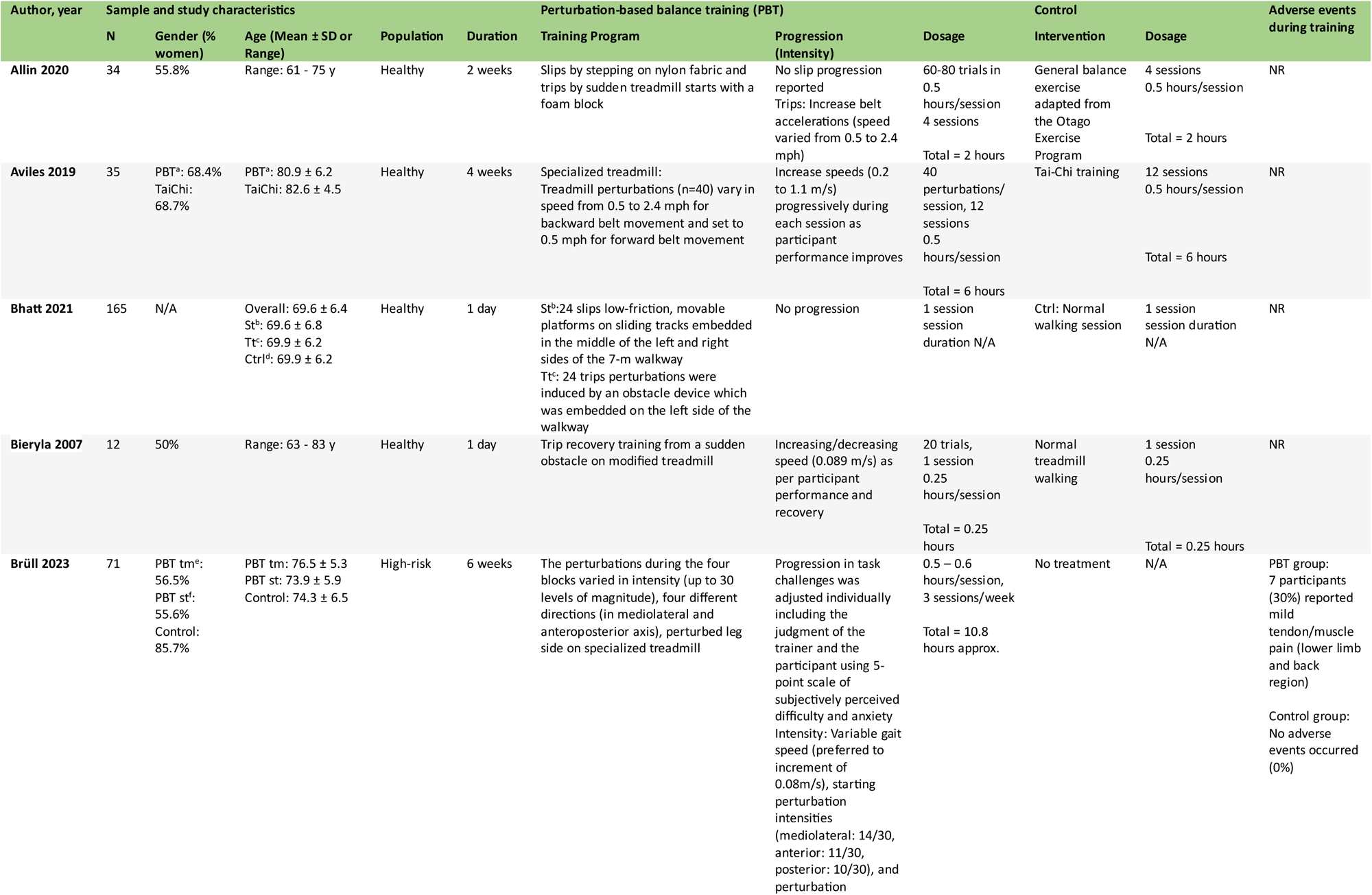
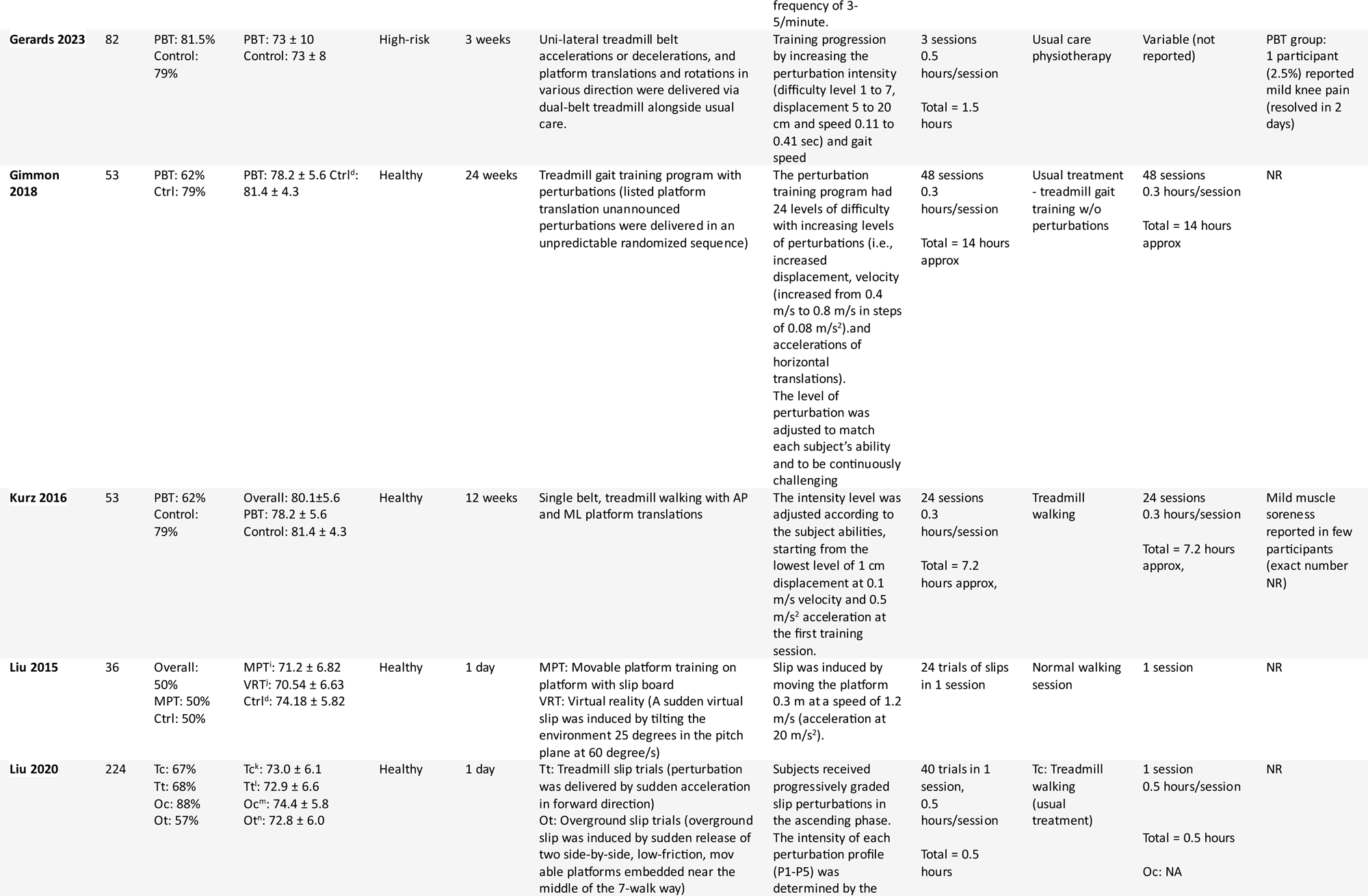
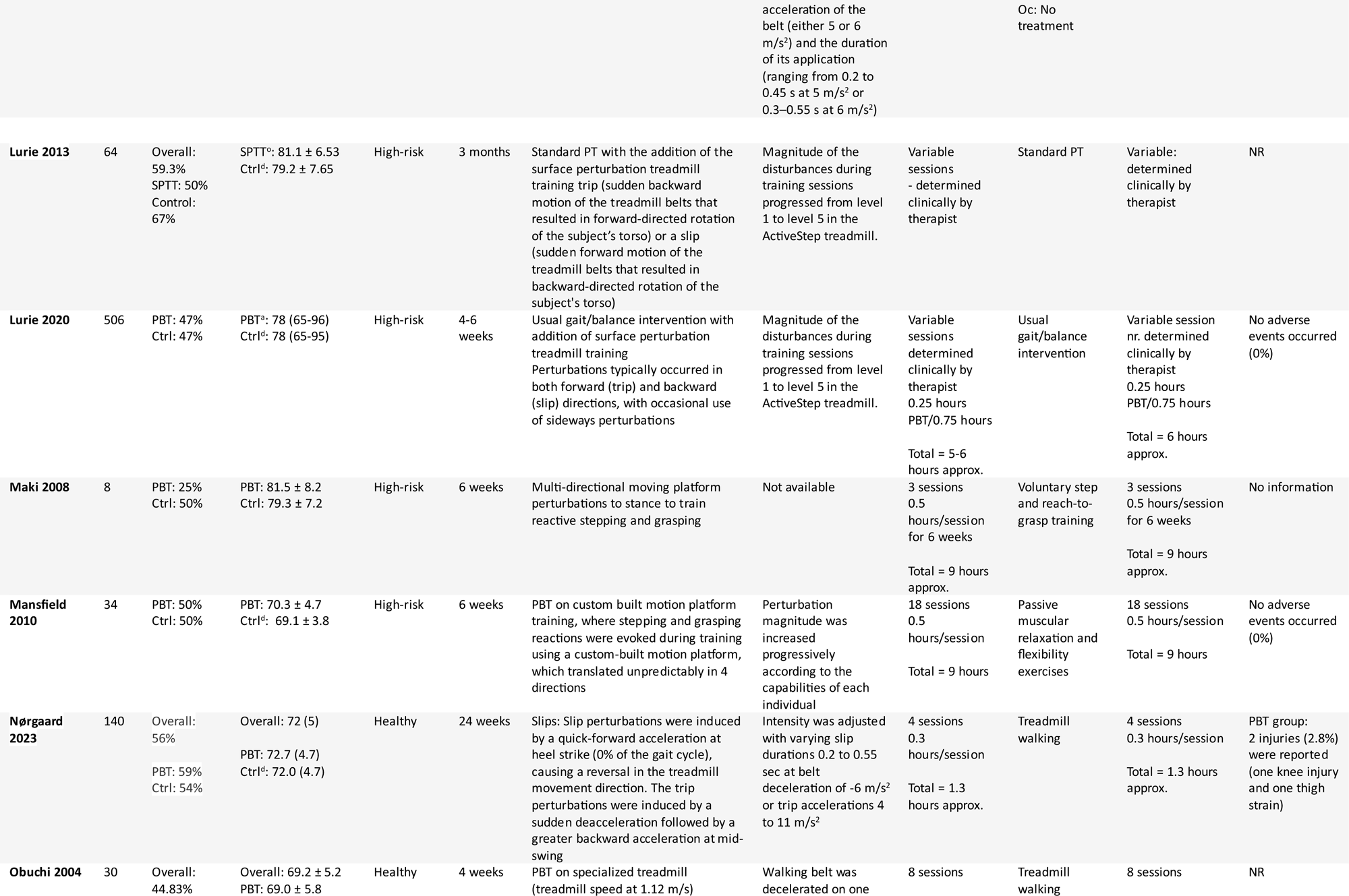
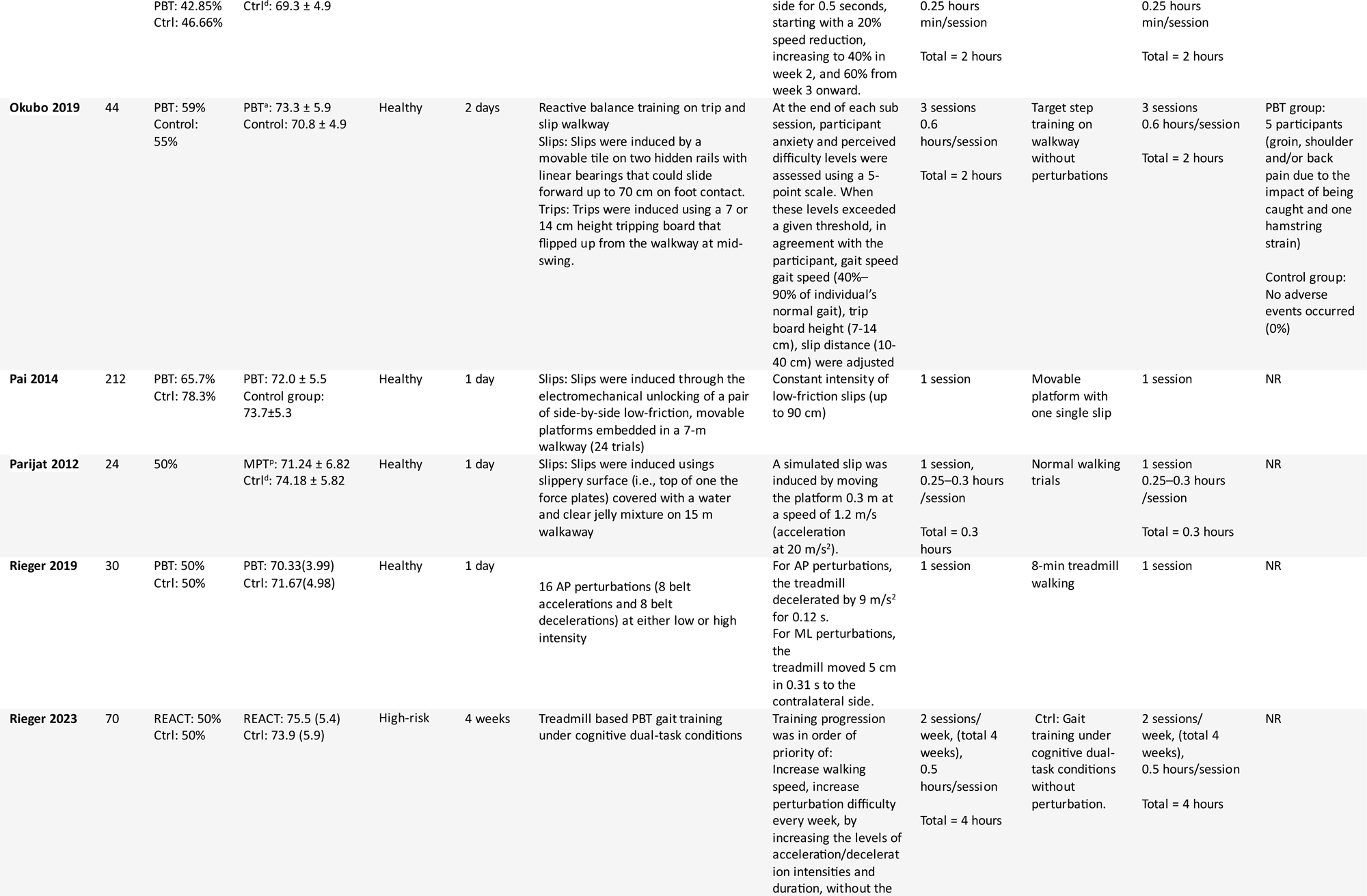
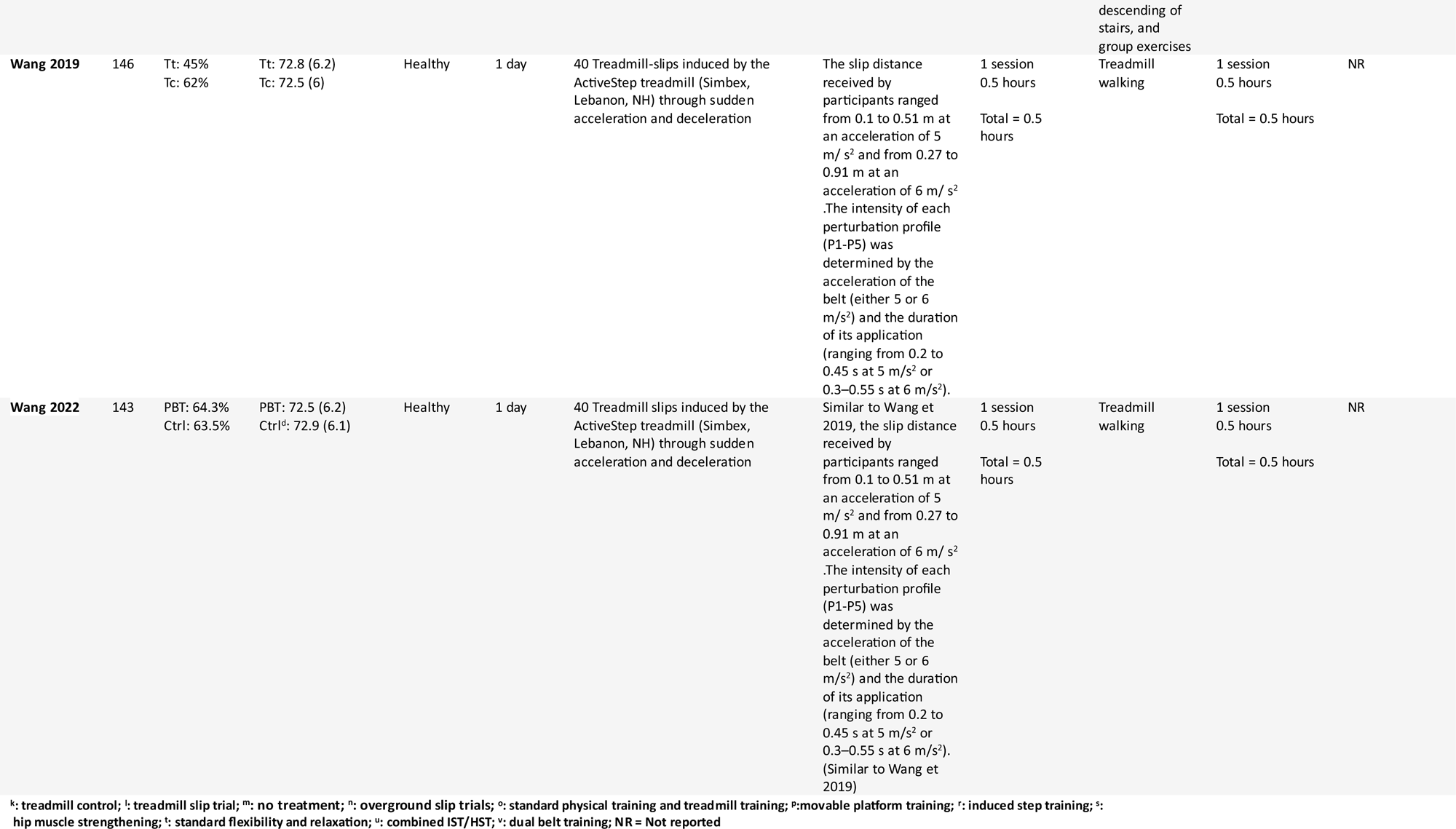
Descriptive characteristics of included studies.

## Supplementary Figures

**Supplementary Figure 1.**
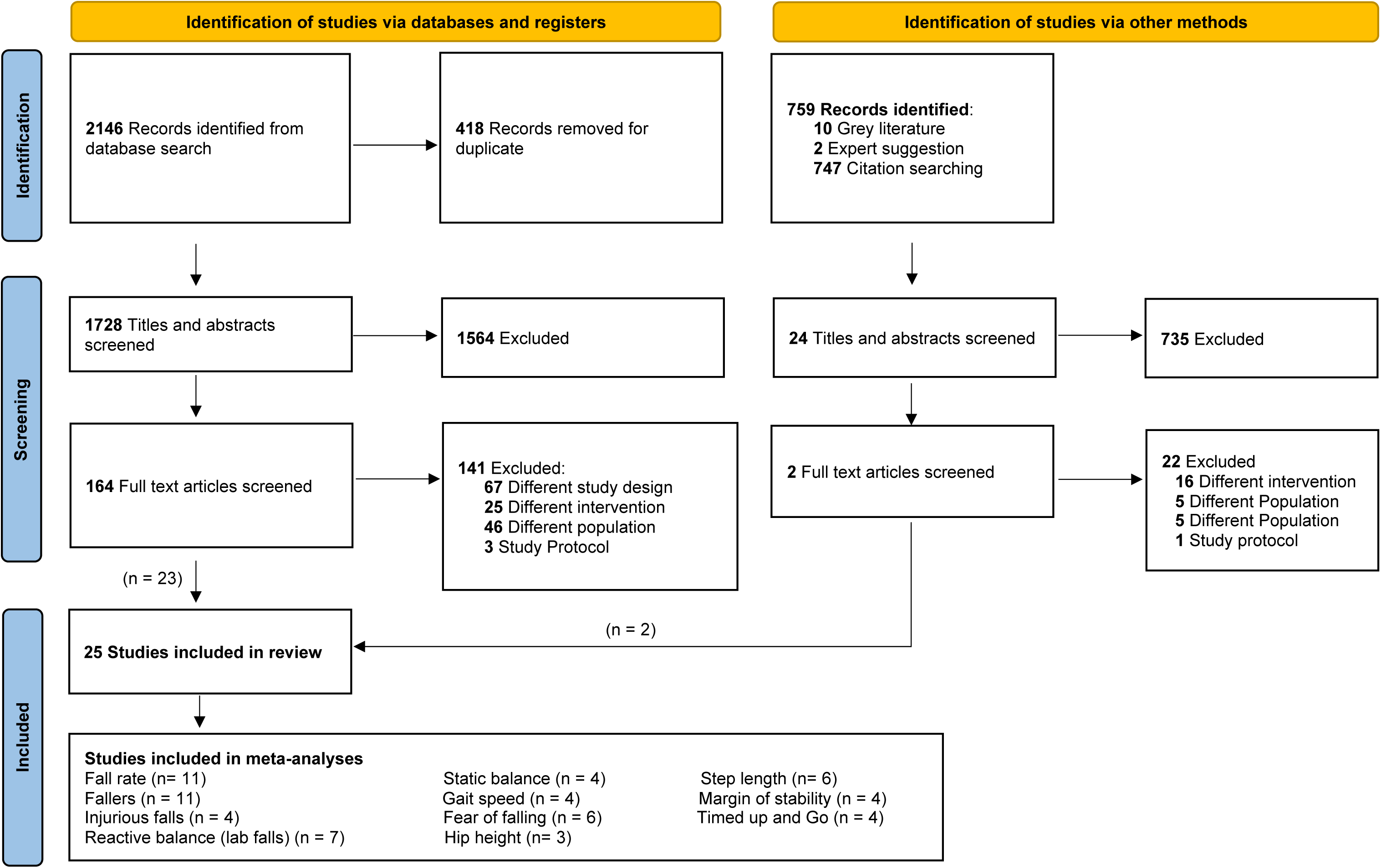
Flow diagram for this systematic review including searches of databases, registers and other sources. Studies that reported the outcome of interest were included in the meta-analysis.

**Sup Figure 2.**
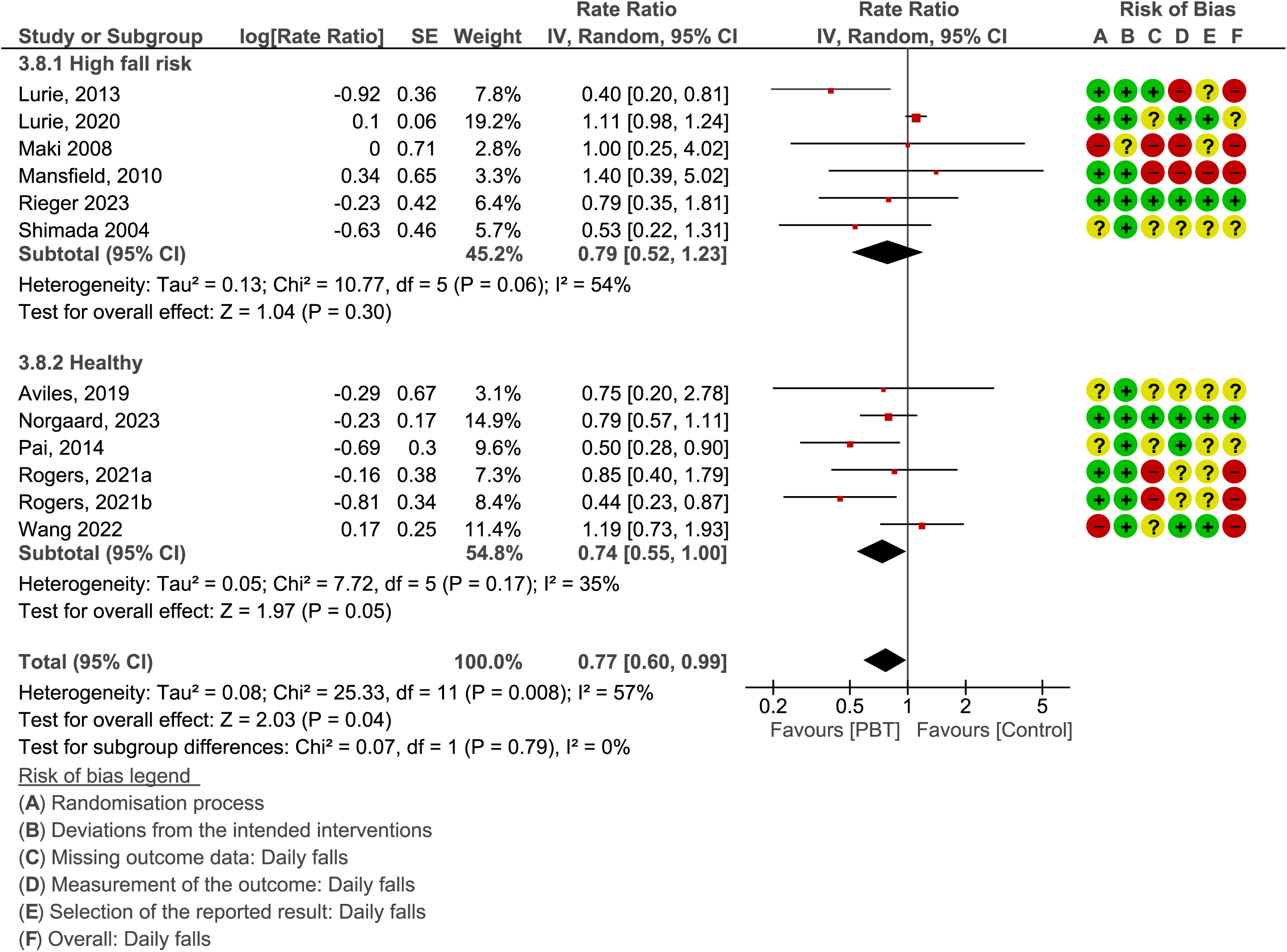
Rate ratio - Subgroup analysis Intervention – high risk/healthy (Rogers 2021a – induced step training plus hip streng hening vs hip strengthening; Rogers 2021b – induced step training vs flexibility and relaxation)

**Sup Figure 3.**
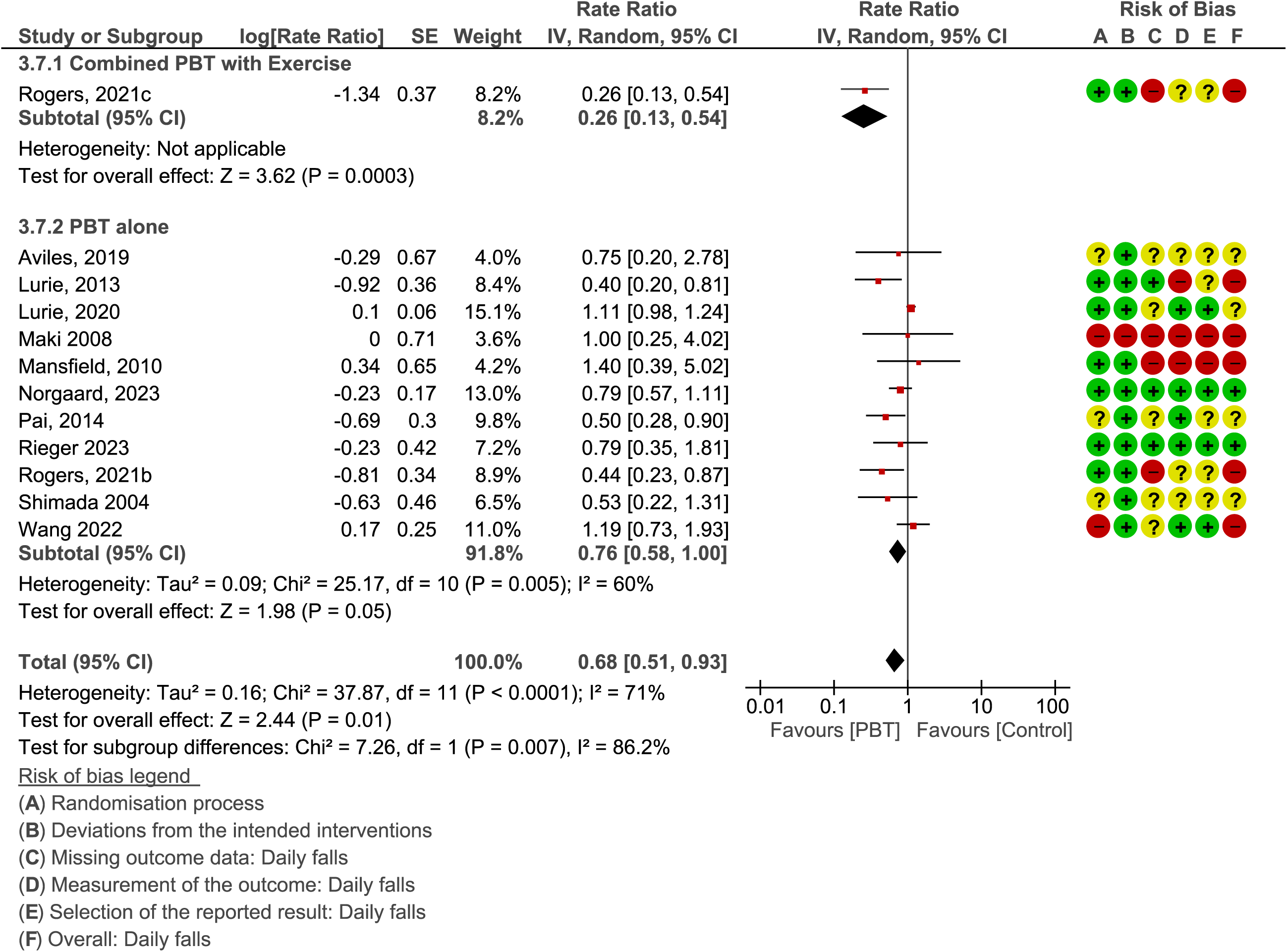
Rate ratio - Subgroup analysis Intervention – Combined PBT with exercise versus PBT alone (Rogers 2021c – induced step training plus hip strengthening vs flexibility and relaxation)

**Sup Figure 4.**
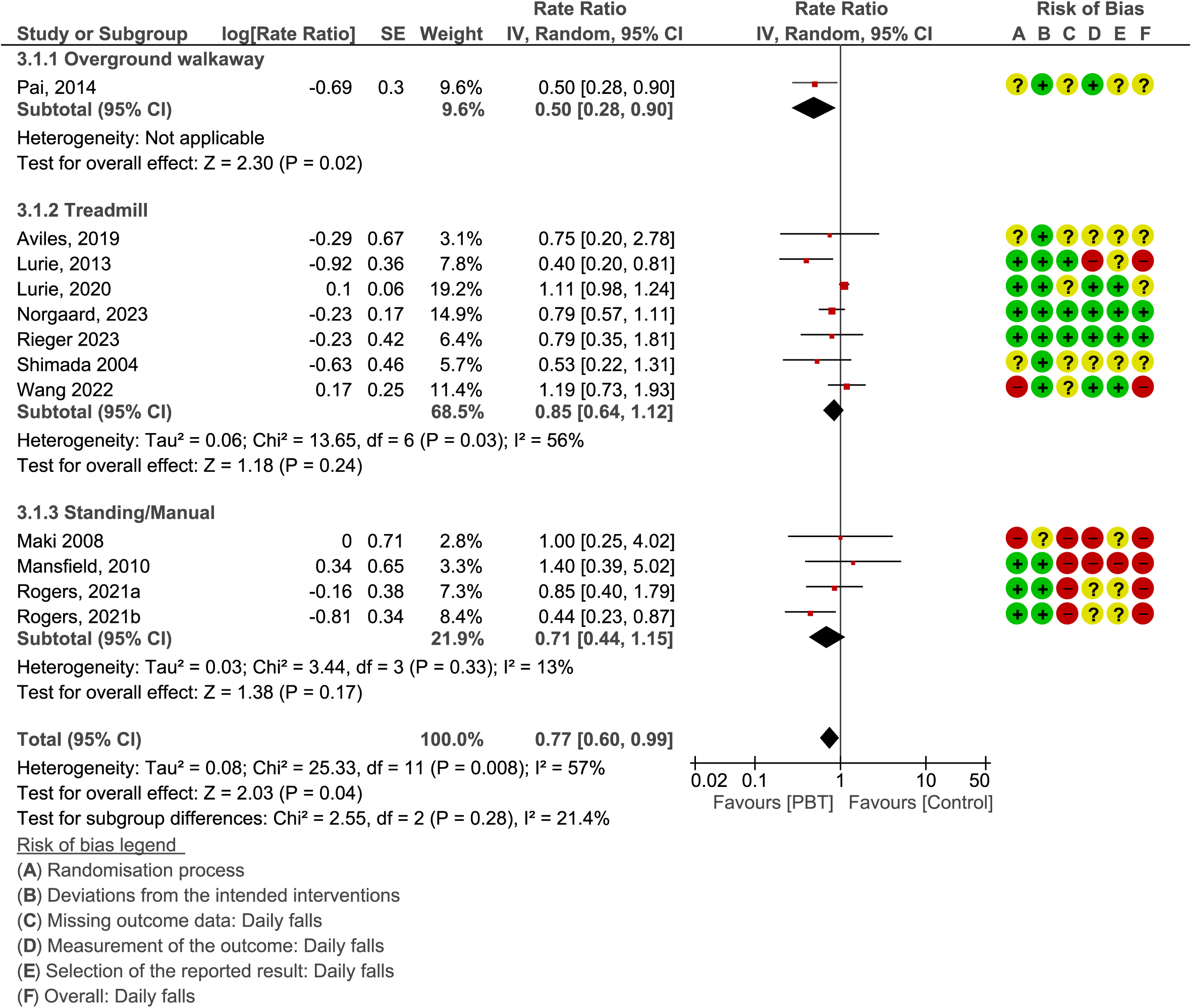
Rate ratio - Subgroup analysis Intervention type 2021a – induced step training plus hip strengthening vs hip strengthening; Rogers 2021b – induced step training vs flexibility and relaxation

**Sup Figure 5.**
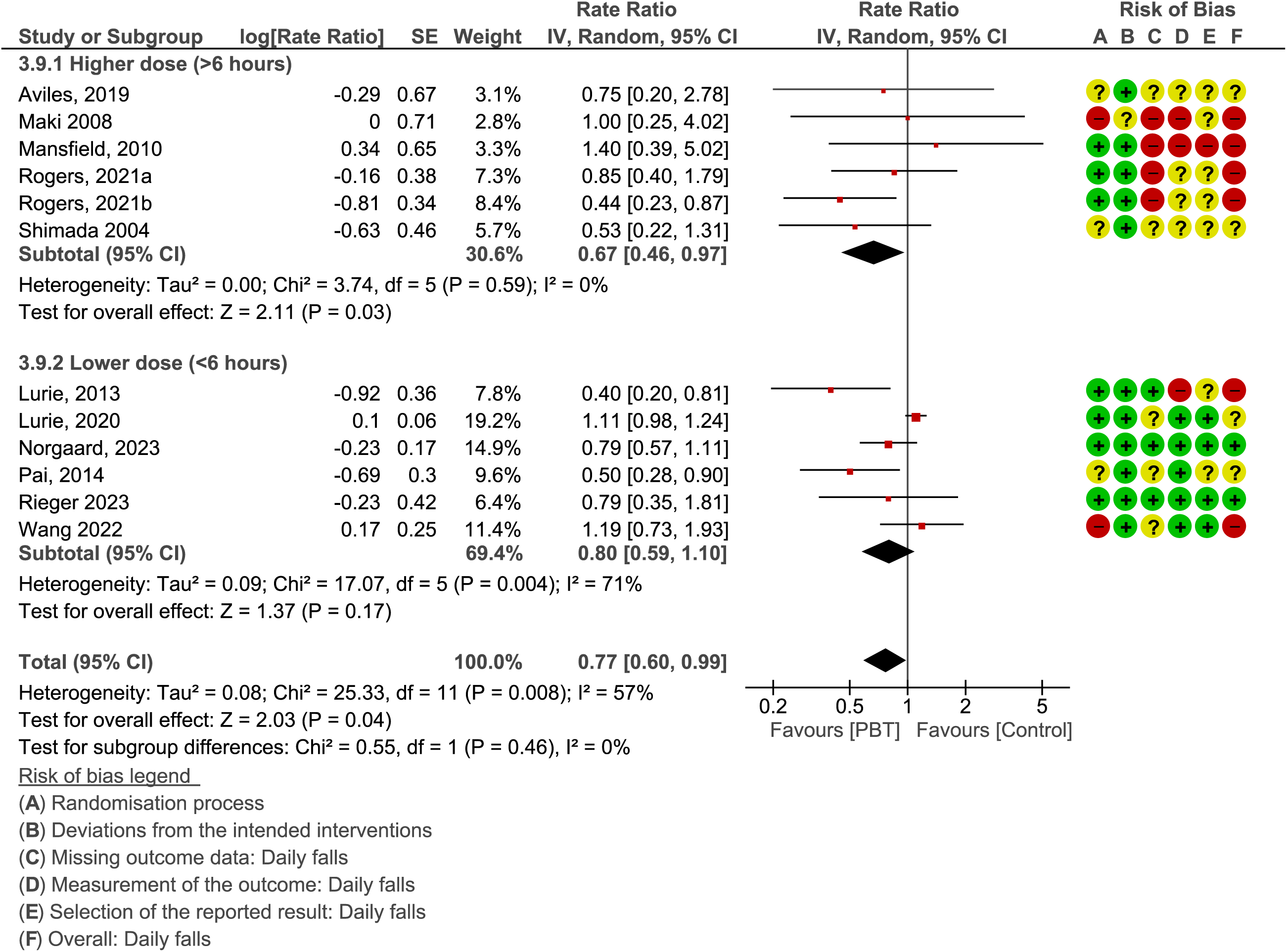
Rate ratio - Subgroup analysis Training dosage (Higher dose versus lower dose) (Rogers 2021a – induced step training plus hip strengthening vs hip strengthening; Rogers 2021b – induced step training and relaxation)

**Sup Figure 6.**
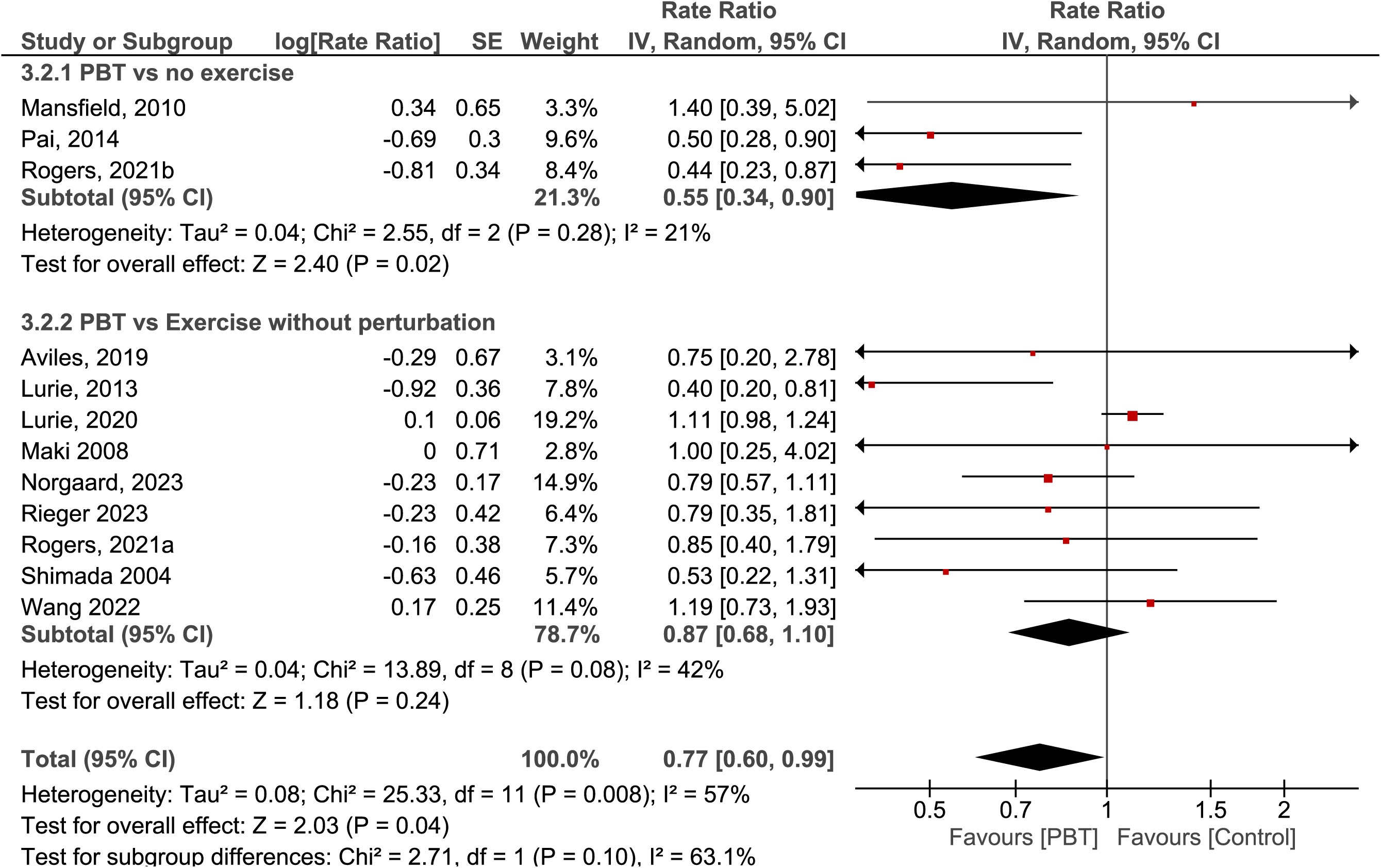
Rate ratio – PBT versus Exercise without perturbation/No exercise (Rogers 2021a – induced step training plus hip strengthening vs hip strengthening; Rogers 2021b – induced step training vs flexibility and relaxation)

**Sup Figure 7.**
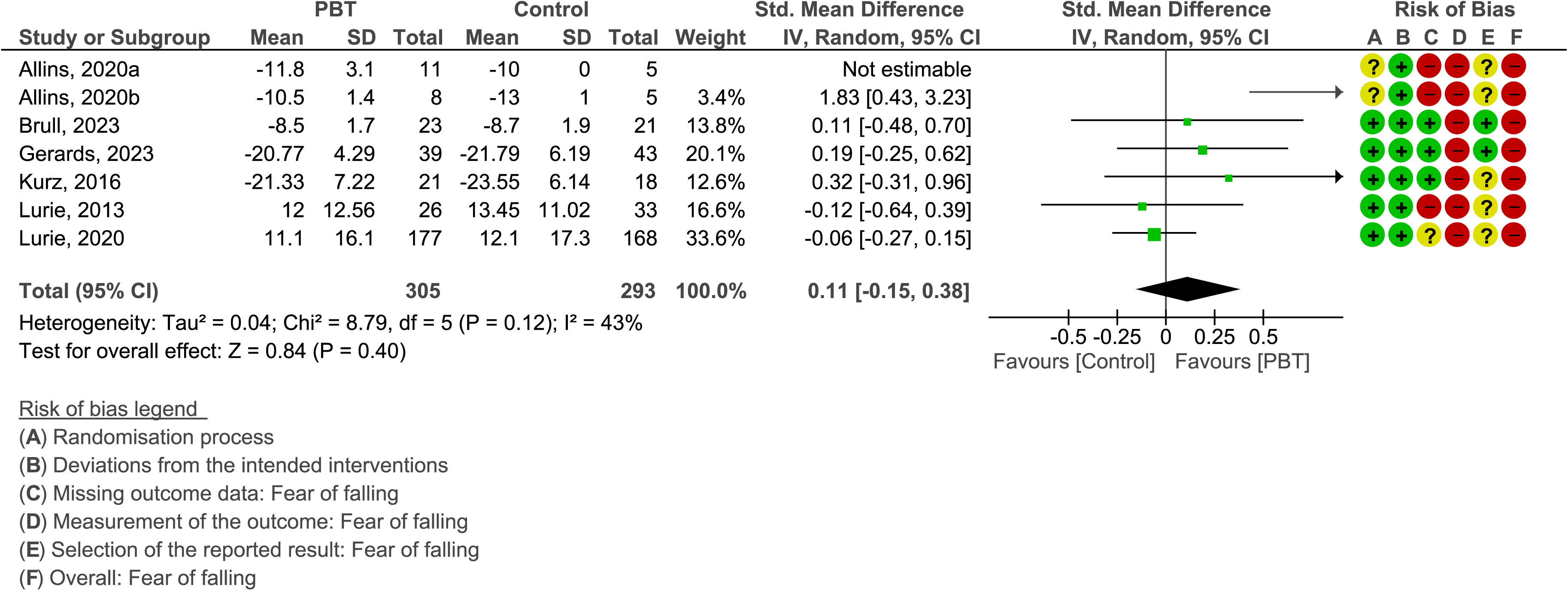
Meta-analyses for Fear of falling 2020a – slip training vs control; Allins 2020b – trip training vs control)

**Sup Figure 8.**
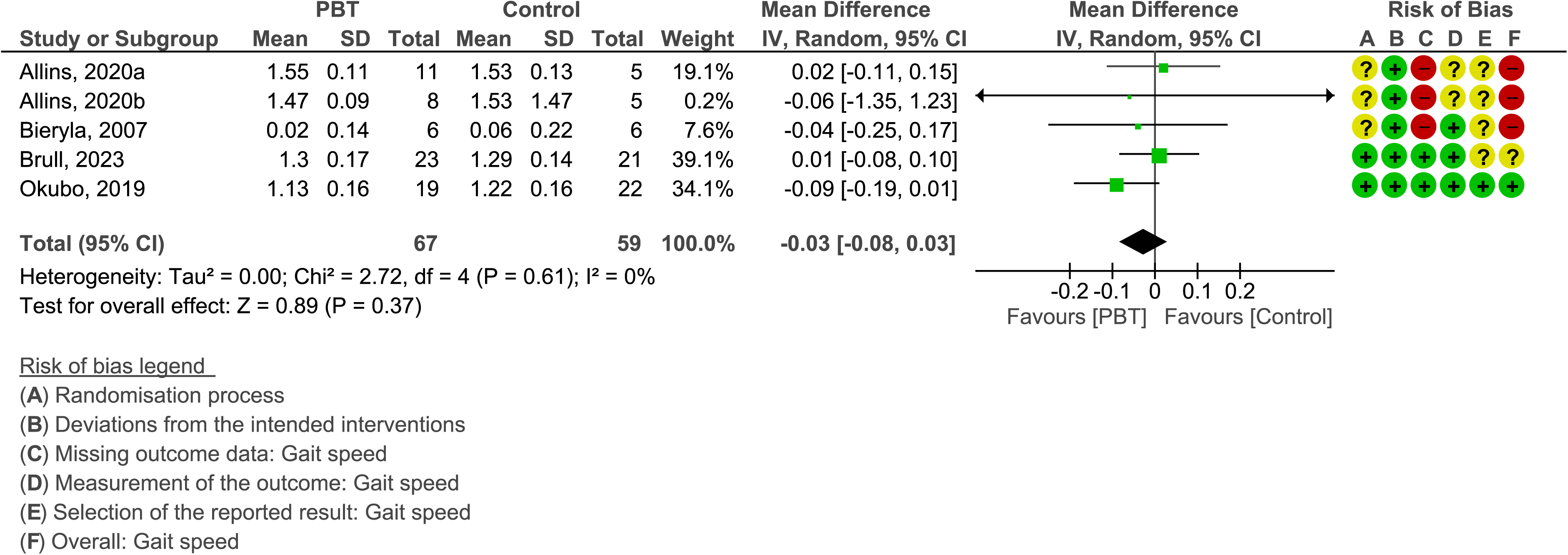
Meta-analyses for Gait speed (m/s) 2020a – slip training vs control; Allins 2020b – trip training vs control)

**Sup Figure 9.**
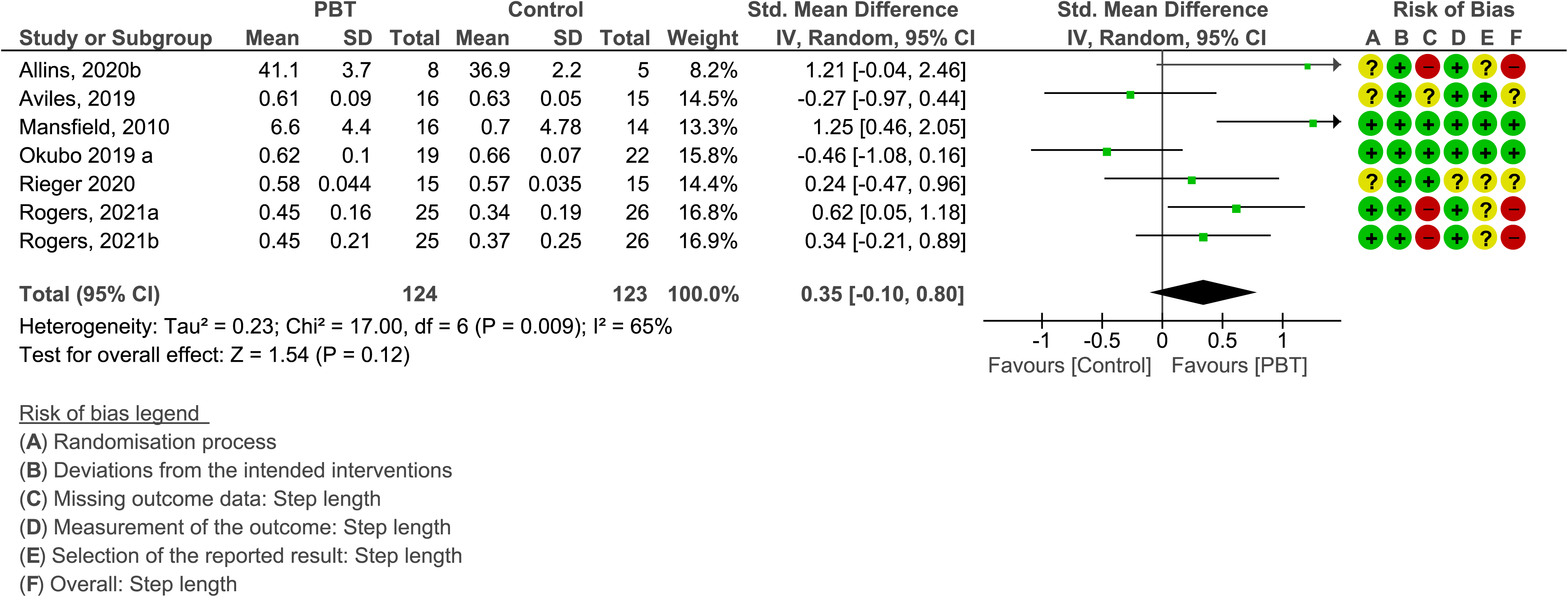
Meta-analyses for Recovery step length (cm) 2020b – trip training vs control; Okubo 2019a – slip training vs control; Rogers 2021a – induced step training plus hip strengthening vs hip training; Rogers 2021b – induced step training vs flexibility and relaxation)

**Sup Figure 10.**
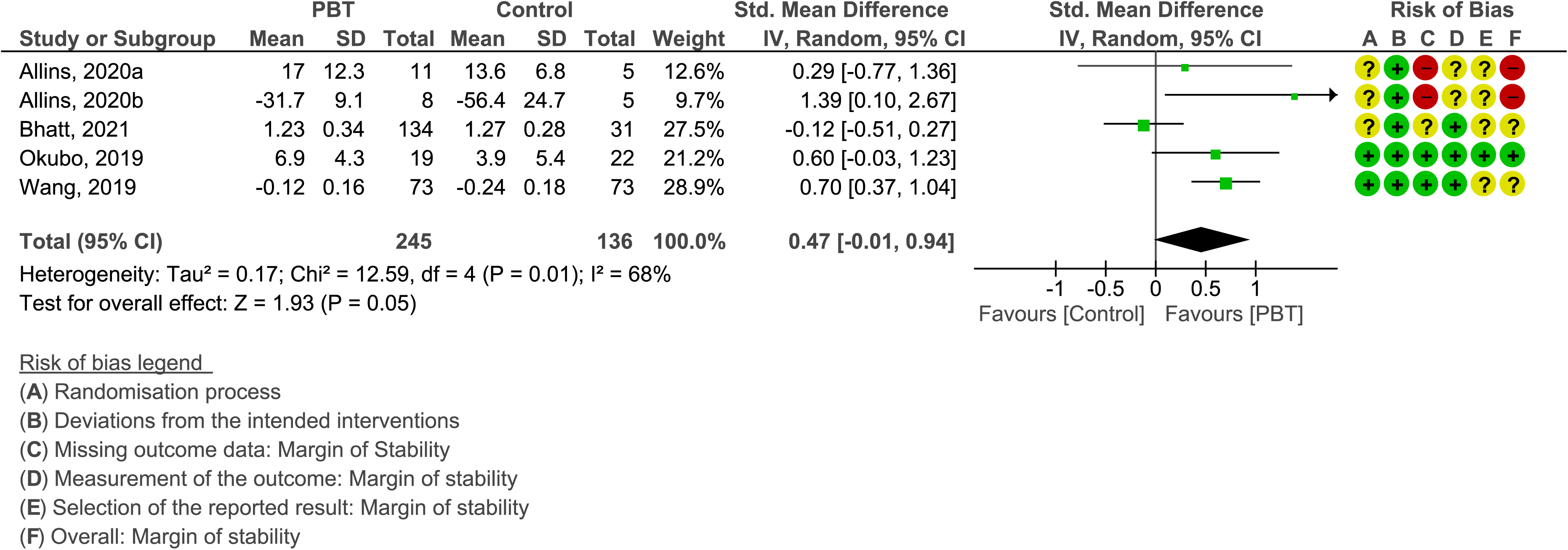
Meta-analyses for Margin of stability during a recovery step (m) 2020a – slip training vs control; Allins 2020b – trip training vs control)

**Sup Figure 11.**
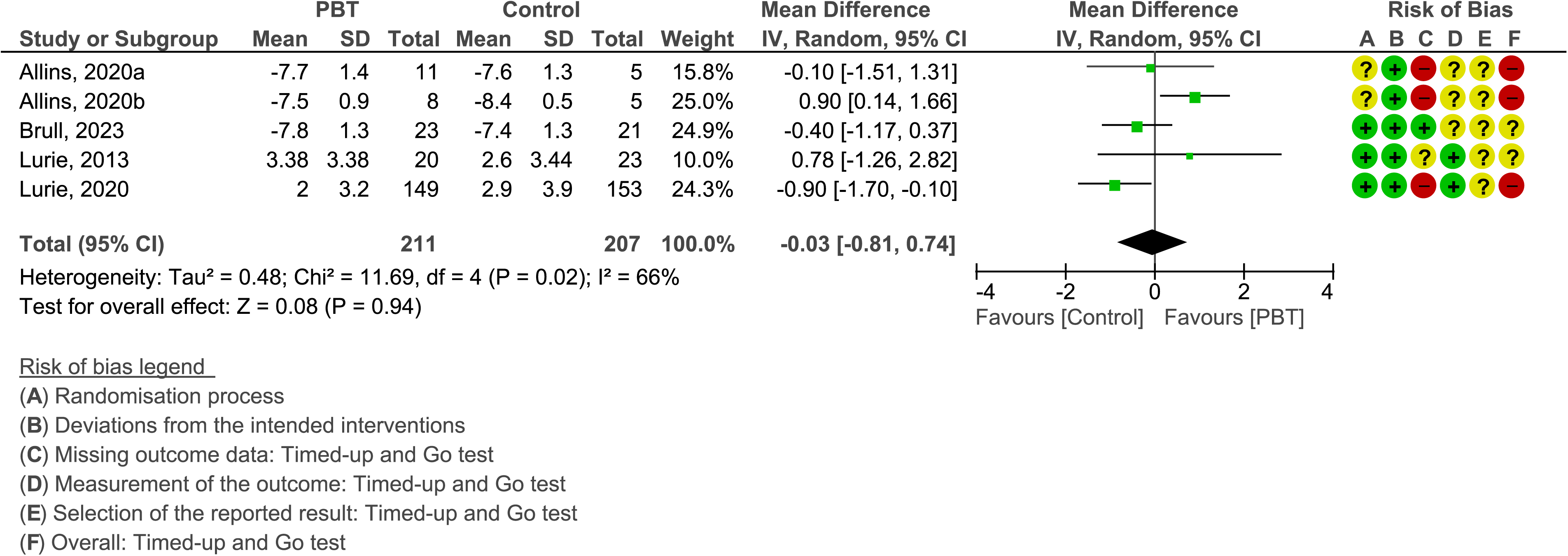
Meta-analyses for the Timed Up & Go test 2020a – slip training vs control; Allins 2020b – trip training vs control)

**Sup Figure 12.**
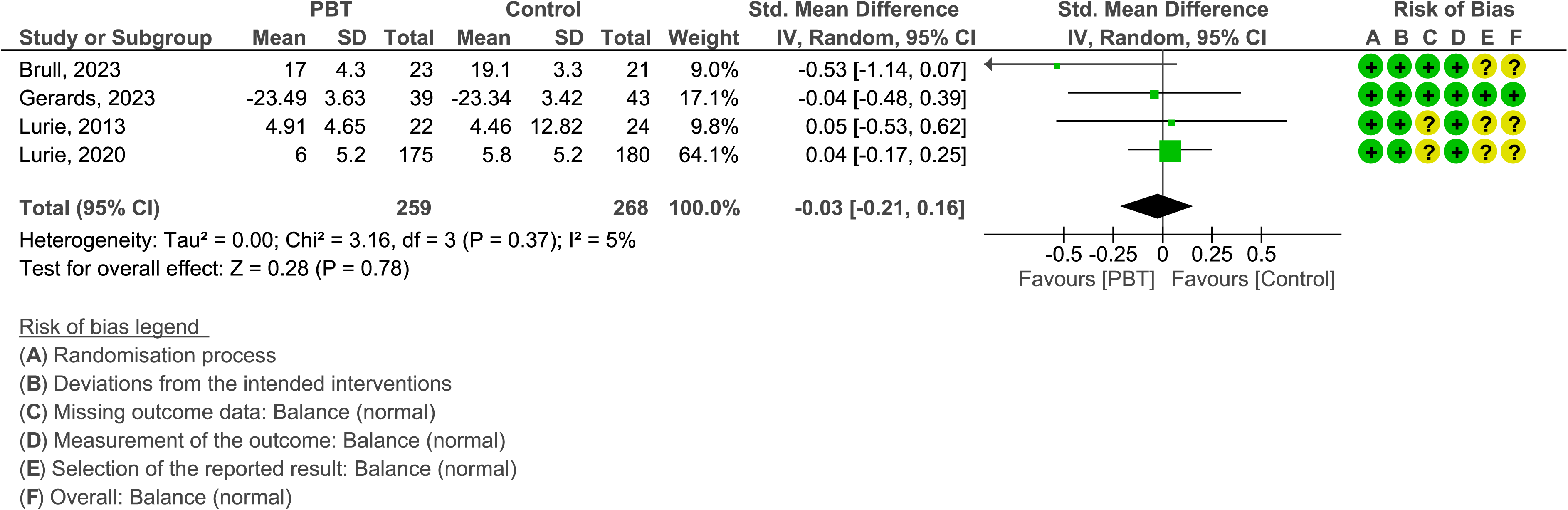
Meta-analyses for Static Balance (normal)

**Sup Figure 13.**
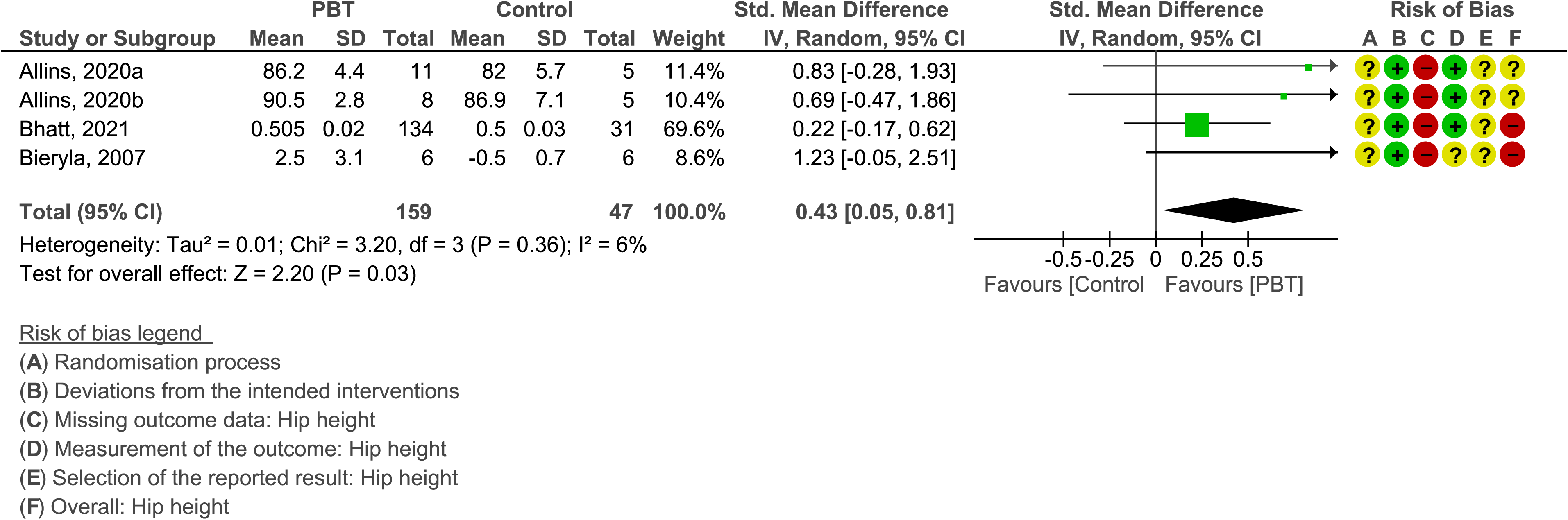
Meta-analyses for Hip height during a recovery step 2020a – slip training vs control; Allins 2020b – trip training vs control)

**Sup Figure 14.**
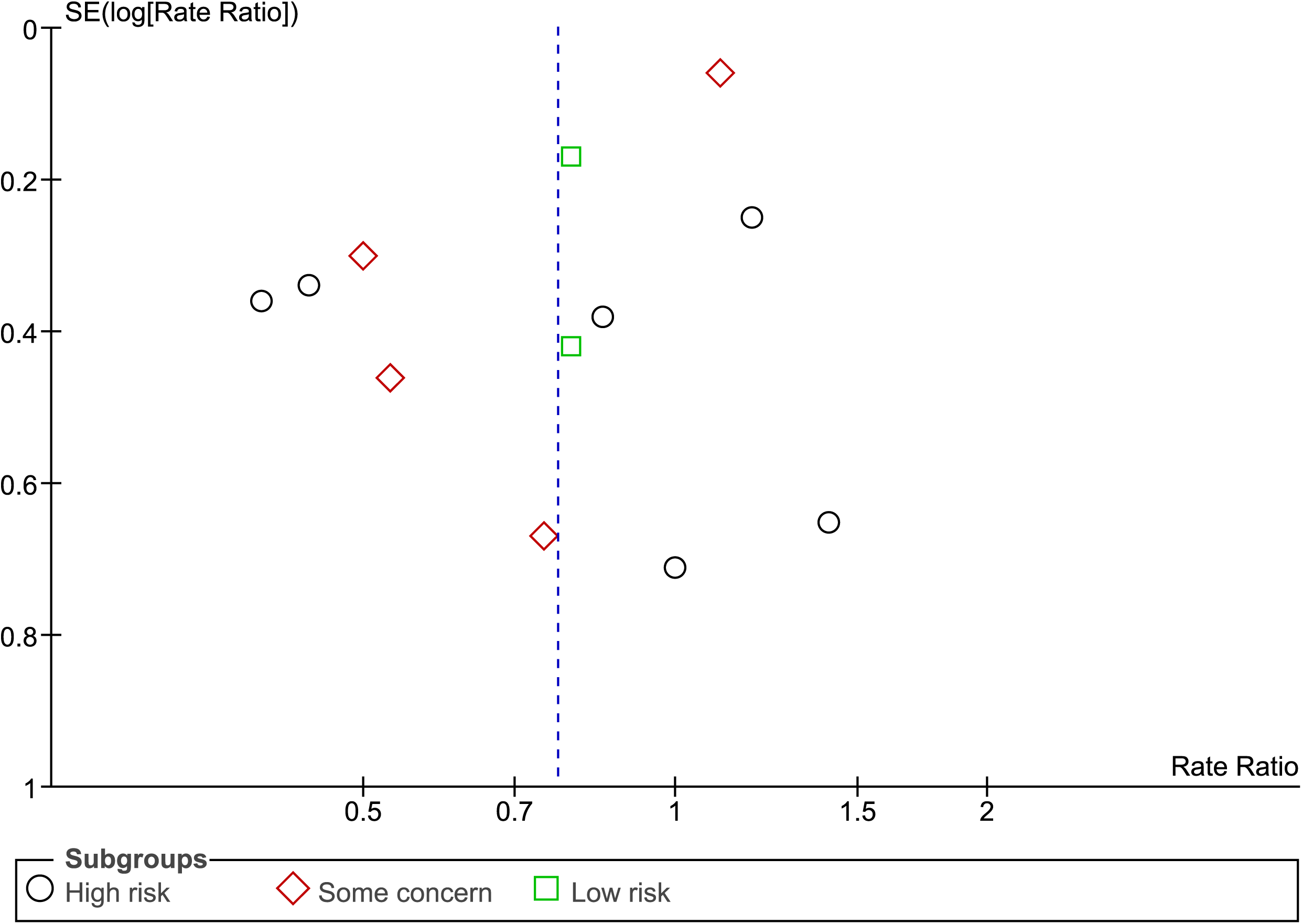
Funnel plot (rate ratio - falls)

**Sup Figure 15.**
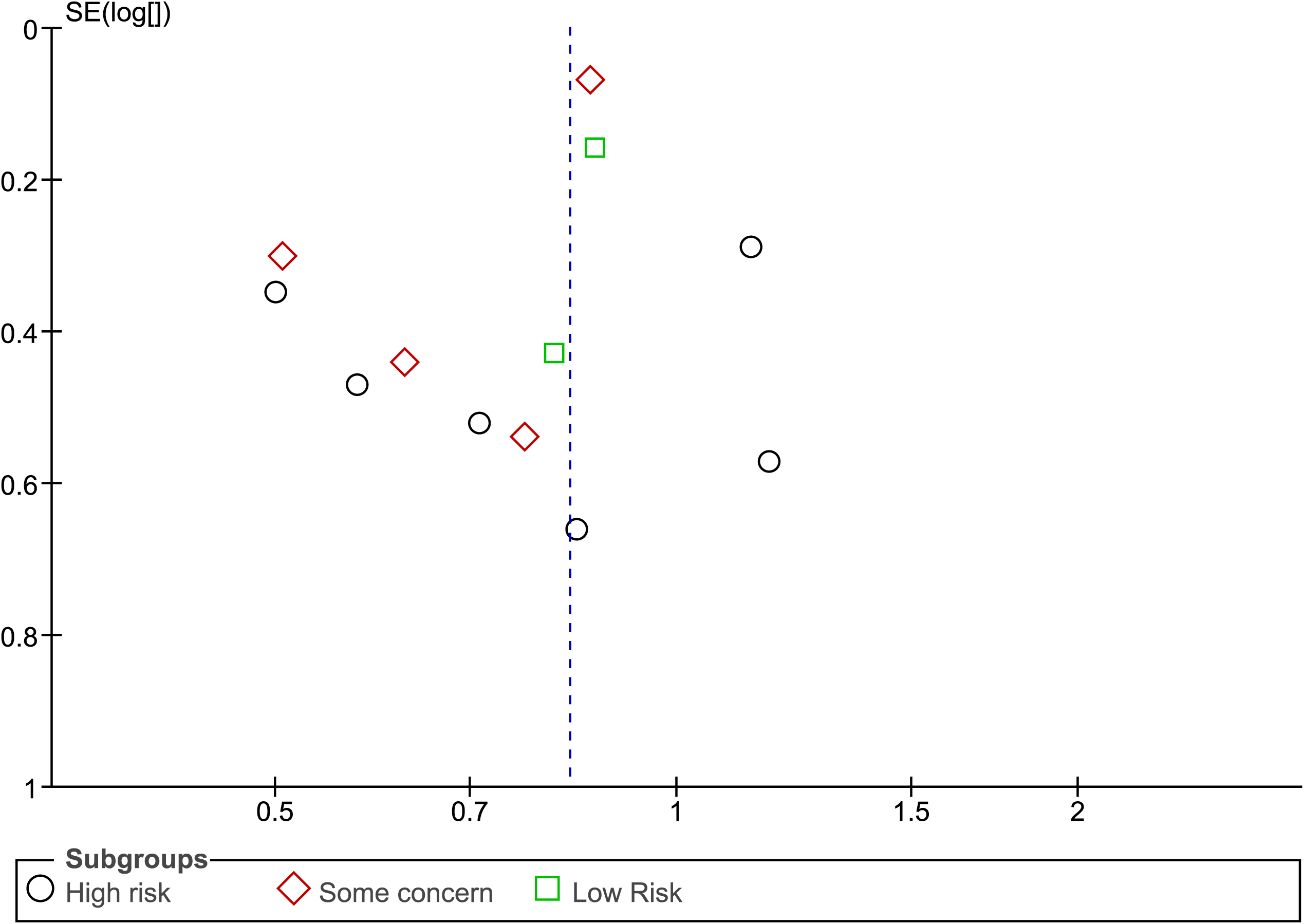
Funnel plot (risk ratio - fallers)

**Sup Figure 16.**
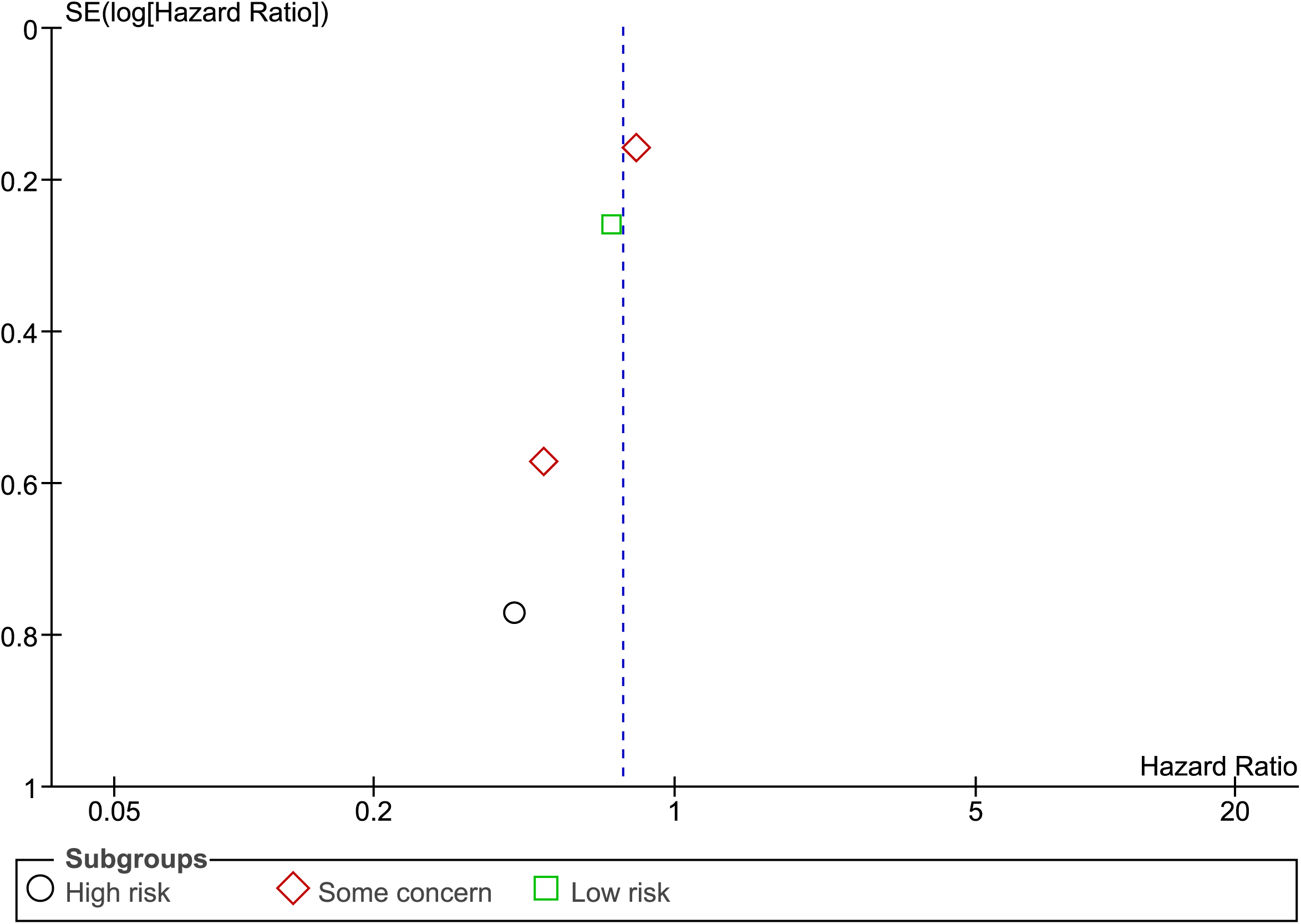
Funnel plot (injurious fallers)

**Sup Figure 17.**
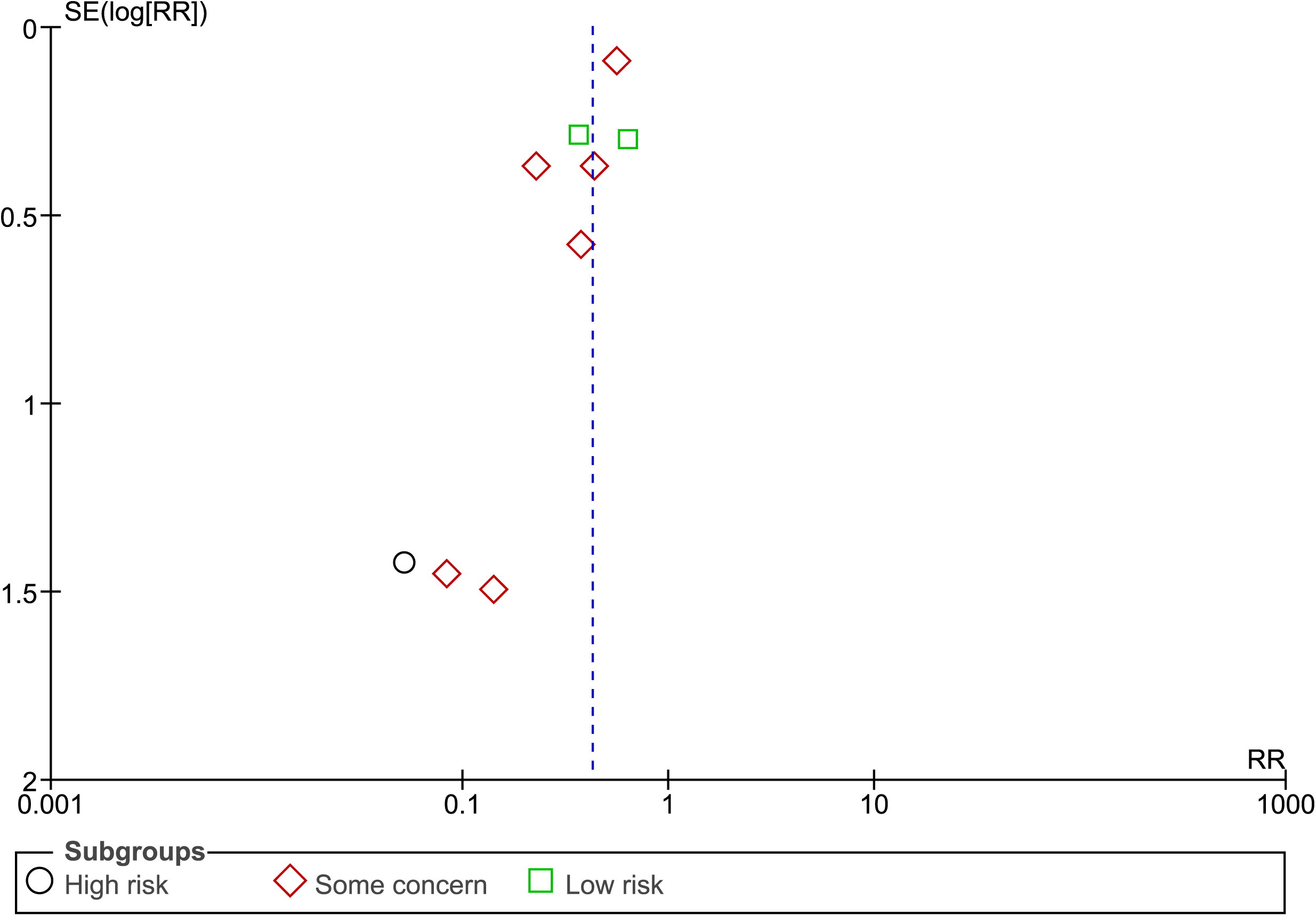
Funnel plot (lab falls)

